# Covariate adjustment for hierarchical outcomes and the win ratio: how to do it and is it worthwhile?

**DOI:** 10.64898/2026.03.30.26347966

**Authors:** Audinga-Dea Hazewinkel, John Gregson, Jonathan W. Bartlett, Samvel B. Gasparyan, David Wright, Stuart Pocock

## Abstract

**Background:** Hierarchical composite outcomes, analyzed using the win ratio, are increasingly used in randomized clinical trials (RCTs). However, methods for covariate adjustment in this context are underdeveloped, despite evidence that adjusting for prognostic variables can increase statistical power.

**Objectives:** Introducing a new covariate adjustment method for hierarchical outcomes using ordinal logistic regression, comparing it with existing approaches, and assessing whether adjustment improves power in randomized trials with hierarchical outcomes.

**Methods:** We developed an ordinal regression-based method for covariate adjustment of the win ratio and compared it with three alternatives: probability index models, inverse probability weighting, and a randomization-based estimator. Methods were applied to the EMPEROR-Preserved rial and tested through extensive simulations involving two common hierarchical outcome structures: time-to-event composites, and composites combining time-to-event with quantitative measures. Simulations assessed impacts on estimates, standard errors, and power across prognostic and non-prognostic settings.

**Results:** In RCT data and simulations, covariate adjustment consistently increased power when adjusting for prognostic baseline variables. Gains were comparable to or greater than those in conventional Cox models, with no power loss for non-prognostic covariates. Our ordinal approach performed similarly to existing methods while providing interpretable covariate effect estimates. Adjusting for baseline values of quantitative components yielded power gains according to the baseline-to-follow-up correlation.

**Conclusions:** Covariate adjustment for prognostic variables meaningfully improves efficiency in win ratio analyses for hierarchical outcomes. Our ordinal method is easily implemented and facilitates covariate effect interpretation. We recommend the broader adoption of covariate adjustment and our ordinal method in randomized trials using hierarchical outcomes.

## 1. Introduction

Composite outcomes are commonly used in the analysis of clinical trials and are often analyzed by considering the time to the first of any event within the composite, for example using a Cox proportional hazards models. However, this approach has limitations. Consider a trial with a composite outcome of death or hospitalization, using the above approach: (1) death and hospitalization receive equal emphasis in the analysis, despite death being clinically more severe; (2) because death can occur after hospitalization but not vice versa, all first hospitalizations count but some deaths are ignored; (3) only the first hospitalization per patient will be captured, whereas multiple hospitalizations may occur. An alternative is to consider a hierarchical composite outcome (HCE) in which clinically more severe outcomes (e.g. death) are prioritized over less severe outcomes (e.g. hospitalization). An added benefit is increased flexibility in terms of the types of outcomes that can be included in a composite. For example, they can combine clinical events with quantitative outcomes such as measures of quality of life, functional capacity or biomarkers. However, in HCEs the target estimand is no longer a specific event rate but a measure of total disease burden. Selection of this estimand may require careful discussion if treatment effects are expected to be discordant across components—for example, when a treatment affects hospitalization but not mortality, including death as a component would dilute the treatment effect. HCE outcomes can be analyzed using the Finkelstein Schoenfeld test ^1^ and the win ratio ^2,3^, or equivalent methods such as generalized pairwise comparisons ^4^.

Most applications of HCEs to date have been in randomized clinical trials (RCTs), particularly in cardiovascular disease ^5^, although HCEs are also used in other therapeutic areas, for example in chronic kidney disease ^6,7^ and COVID-19 ^8^. While covariate adjustment is not necessary for making valid inferences in this setting, it is commonly done for other types of outcome mainly because adjustment for prognostic covariates can improve statistical power ^9,10^ or because the conditional effect is of interest. Several methods have been described for covariate adjustment in the analysis of hierarchical outcomes with win ratio or related methods ^11–13^, but there has been no systematic comparison of these methods and, most importantly, there is no clear guidance for when to use covariate adjustment when analyzing HCEs. It is therefore timely to review available methods for covariate adjustment for hierarchical outcomes and establish to what degree adjustment for prognostic covariates could improve statistical power in RCTs. In addition, we introduce a new method for covariate adjustment with hierarchical outcomes and compare it with other methods. While existing methods can quantify the effect of covariates when estimating the win odds, they cannot do so for the win ratio. Our proposed ordinal win ratio estimator provides a conditional estimate of the treatment effect and interpretable covariate effect estimates, which is analogous to conventional covariate adjustment in models like the Cox model.

The aims of this paper are therefore (1) to introduce our new approach to covariate adjustment for hierarchical outcome; (2) to review existing methods and compare them to ours; (3) to see how much covariate adjustment improves statistical power with hierarchical outcomes. To do this, we perform analyses with and without covariate adjustment in a large randomized controlled trial ^14^ and simulation studies.

## 2. Hierarchical outcomes and the win ratio

Hierarchical composite outcomes work by selecting the outcomes, defining their prioritization, and comparing pairs of patients, one from the intervention arm and one from the control arm based on their events and the defined prioritization. Within each pair, one compares their outcomes to determine which patient had a better outcome. Consider a hierarchical outcome of death (highest priority) and hospitalization. One begins by comparing patients based on death (during their shared follow-up, in variable follow-up designs). If the intervention patient survives longer than the control patient it is a ‘win’ because the better outcome occurs in the intervention arm, and if the control patient survives longer, it is a ‘loss’. Once a win or loss is determined, outcomes lower in the hierarchy are not considered. But for patient pairs who are tied based on death the next level of the hierarchy (hospitalization) is considered: patients are compared with regards to time to first hospitalization, so that if the intervention (control) patient was hospitalized first it would be a ‘loss’ (‘win’), respectively. Patient pairs that cannot be separated at any level of the hierarchy are a ‘tie’. Across all patient pairs, one divides the total number of wins by the total number of losses to estimate the win ratio.

The most common application ^5^ is the win ratio which has the same underlying methodology as generalized pairwise comparisons ^4^. With N_I_ intervention patients and N_C_ control patients, all N_I_×N_C_ possible patients pairs are compared. The win ratio is defined as 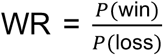, where *P*(win) is the probability of a win in the intervention group over the control group, and *P*(loss) the probability of a loss in the intervention group, with e.g., 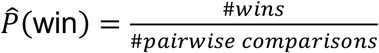, where #*pairwise comparisons* = *N*_*I*_ × *N*_*C*_ The win ratio is calculated as 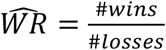. There are available statistical methods to construct the 95% confidence intervals and p-values (implemented in software program R and Stata). The win ratio can be interpreted as the odds that for a randomly chosen pair of patients that are not tied, the better outcome occurs in the intervention patient. Alternative measures of treatment benefit for hierarchical outcomes include the win difference, also known as the net clinical benefit, defined as *WD* = 100[*P*(win) − *P*(loss)], typically estimated as 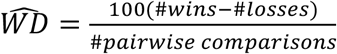; the win odds, defined as 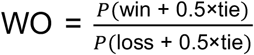 and estimated as 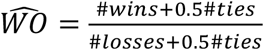; and the Mann-Whitney probability *θ* = *P*(win + 0.5×tie), estimated as 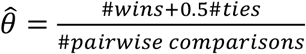. Note that the net clinical benefit can also be calculated from the Mann-Whitney probability as *WD* = 100[2 × *θ* − 1] and the number needed to treat as 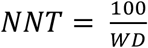, rounded up. In scenarios where there are no ties, typically when a continuous quantitative outcome is included in the hierarchy, the win odds and win ratio are the same.

## 3. Ordinal logistic covariate adjustment for the win ratio

We propose a method for covariate adjustment for the win ratio which uses ordinal logistic regression. The approach works by applying ordinal logistic regression to pairwise comparisons (as opposed to the ordinary application to patient-level data) as follows:

1. Form pairs of patients across trial arms, so that each patient i from the intervention group is paired with each patient *j* from the control group, so that there are *N*_*I*_ × *N*_*C*_ paired comparisons. For each paired comparison, generate a response variable *Y*_*ij*_ which takes the value *Y*_*ij*_=0 if patient *j* has a better outcome than patient *i* (a ‘loss’ for the intervention group), *Y*_*ij*_ = 1 for a tie and *Y*_*ij*_ = 2 if patient *i* has a better outcome than patient *j* (a ‘win’ for the intervention group).
2. Let Δ*C* denote within-pair difference in covariate values, i.e. Δ*C*_*ij*_ = *C*_*i*_ − *C*_*j*_.
3. An ordinal logistic regression model ^15^ is then fit to the resulting data and modelled as:

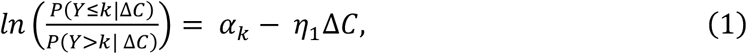

f*or k* = 0 *or* 1. In this model *α*_0_ = *logit*(*P*(*Y* ≤ 0 | Δ*C* = 0)) so that 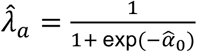 is the estimated adjusted loss probability (i.e. the estimated loss probability for two patients with the same covariate value). Likewise, 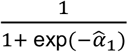 represents the estimated probability of either a loss or a tie. It follows that 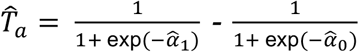 is the estimated adjusted probability of a tie and 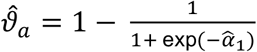 is the estimated adjusted win probability. *η*_1_ represents the log common odds ratio for a one unit increase in covariate difference in the covariate value ΔC between the intervention patient and control patient. Note that while the unadjusted win ratio does not require any assumptions, here we make the proportional odds assumption for the covariate, i.e. the odds ratio of the covariate on moving from a loss to a tie or win is the same as when moving from a loss or tie to a win. The adjusted win ratio, win odds and win difference can each be calculated using the relevant adjusted probabilities of a win, loss or tie. For example, the win ratio is calculated as 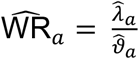 and win difference as 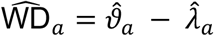. The formula in (1) can be extended to multiple covariates, each with its own coefficient. Confidence intervals and p-values for the adjusted win ratio can be calculated by bootstrapping, or alternatively by using a formula for the standard error based on the delta method, as derived in Appendix A. This same appendix provides a worked example for estimating the ordinal adjusted WR with SE in *R*.

Note that our adjusted win ratio estimates the win ratio for two patients conditional on identical covariate values (conditional treatment effect), whereas the unadjusted win ratio estimates a marginal win ratio (average treatment effect) across all pairs of patients irrespective of covariate values. Due to non-collapsibility of win ratio, the conditional win ratio targets a different estimand than the unadjusted win ratio ^16,17^. Like other conditional models, such as logistic regression or the Cox model, our model is not robust to model misspecification. Specifically, the proportional odds assumption may be violated if there is effect modification by one of the covariates (i.e. if treatment effect varies by covariate level). Note that the type-1 error is still expected to be controlled in the RCT setting (Section 6.1 and Appendix A.2).

## 4. Existing methods for covariate adjustment with hierarchical outcomes

### 4.1. Probability index models

Conceptually most similar to our ordinal logistic regression based approach is to use probability index models ^12^. This method estimates a conditional win odds, which can be estimated using a logistic regression with a win/tie/loss indicator for each patient pair as the dependent variable and pairwise differences in treatment indicator and covariate values as the independent variables. The resulting treatment coefficient from the logistic regression is a log odds of a win for two patients with the same covariate value, a conditional estimand. A drawback of this approach is that although it can estimate the win odds and the Mann-Whitney probability, it cannot be used to calculate the win ratio. Furthermore, while the Thas probability index model was specified for a single patient-level ordinal outcome, we apply it to pairwise comparisons, where the outcome (win, tie, or loss) is generated with respect to all components of the hierarchical composite outcome.

The method works as follows:

1. Form all possible non-redundant pairs of patients, including comparisons between patients in the same arm so that there are 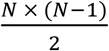 paired comparisons, with *N* = (*N*_*I*_ + *N*_*C*_). For each paired comparison, generate a response variable *W*_*ij*_, which takes the value *W*_*ij*_=0 if patient *j* has a better outcome than patient *i, W*_*ij*_ = 0.5 for a tie and *W*_*ij*_ = 1 if patient *i* has a better outcome than patient *j*. The indices for the comparisons are defined as 1 ≤ *i* < *j* ≤ *N*, to avoid redundant comparisons (e.g., *W*_12_ is defined, comparing patient 1 to patient 2, but not the redundant *W*_21_, comparing patient 2 to patient 1).
2. Let Δ*X* denote a vector of differences in the treatment indicator (*X*_*ij*_ = 0 if both patients are in the same group, *X*_*ij*_ = 1 if patient *i* is in the intervention group and *j* is in the control group and *X*_*ij*_ = −1 if patient j is in the intervention group and *i* is in the control group.
3. Let Δ*C* denote within-pair difference in covariate values, i.e. Δ*C*_*ij*_ = *C*_*i*_ − *C*_*j*_.
4. Then the following regression is performed:

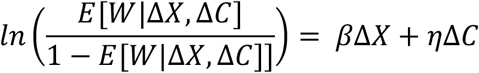

This logistic regression estimates a treatment effect *β* and the adjusted win odds is then calculated as WO_*a,glm*_ = *e*^*β*^. Vermeulen *et al*. ^18^ later developed a further extension to this model which integrates over covariate values and estimates a marginal covariate-adjusted Mann-Whitney probability (and win odds). We consider two alternative marginal estimators later in sections 4.2 and 4.3.

Both the Thas ^12^ and Vermeulen ^18^ papers frame their results in terms of the generalized pairwise comparisons framework and the Mann-Whitney probability, with a primary focus on non-hierarchical continuous or ordinal outcomes. The statistical implementation in the *pim* R package ^19^ follows the theory presented in Thas *et al*. for a single-level ordinal outcome, and does not accommodate either a time-to-event outcome or a multi-level hierarchical outcome, while we apply it to pairwise comparisons with no restrictions on outcome type. A recent preprint ^20^ extends Thas’ and Vermeulen’s work by re-expressing the covariate-adjusted Mann-Whitney probability as the win odds, describing as method to obtain covariate-adjusted win odds for a multi-level hierarchical outcome.

### 4.2. Randomisation-based method

The randomization-based (RB) method was first described by Koch ^*21*^, and later framed by Gasparyan in terms of the win odds ^11^. Similar to probability index models, this method focusses on calculating an adjusted win odds, rather than a win ratio. The process begins with estimating the (unadjusted) Mann-Whitney probability *P*(*w*i*n*) + 0.5P(*tie*) using the intuitive estimator 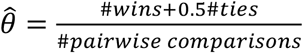. To adjust for covariates, an adjustment term is subtracted from 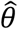 yielding the estimator:

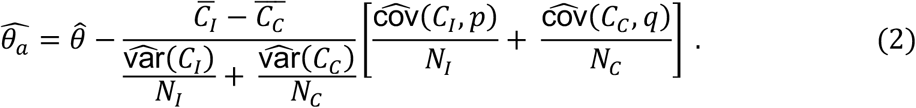

Here:

- *C*_*I*_ and *C*_*C*_ are covariate values for patients in the intervention and control groups respectively
- 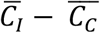 is the difference in their groups means
- *N*_*I*_ and *N*_*C*_ are sample sizes in the intervention and control groups, respectively
- *p* is a vector of length *N*_*I*_ where each element *p*_*i*_ is the average win + 0.5 tie indicator for patient *i* in the intervention group, that is 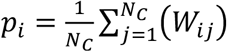
- *q* is a vector of length *N*_*C*_, where each *q*_i_ is the average loss + 0.5 tie indicator for patient *j* in the control arm: 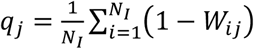, where W is an indicator variable denoting win/loss/tie status, with *W*_*ij*_ = 1 if the *i*th patient from the intervention group wins against the *j*th patient from the control group, *W*_*ij*_ = 0 if the converse is true, and *W*_*ij*_ = 0.5 if the two patients are tied.

To give some intuition as to why this adjustment works, we note that 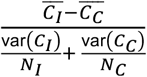 represents a standardized mean difference between groups in the average covariate value, while 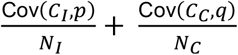 measures how strongly the covariate is associated with the win/tie/loss indicator, with the latter term allowing for the quantification of the prognostic value of the covariate. The adjustment is performed for each covariate separately. When multiple covariates are present, the calculation of each adjustment term is slightly different, and accounts for correlations amongst covariates before the term is subtracted from 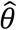. More details on this are provided in Appendix B. Once the adjusted estimator, 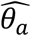, is obtained, the adjusted win odds can be calculated as 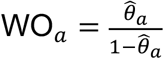. In Koch’s and Gasparyan’s original papers and subsequent statistical software ^22,23^, this method is implemented for estimating the adjusted Mann-Whitney probability and win odds when adjusting for multiple covariates, and for univariate HCEs (multiple outcomes collapsed into a single outcome per patient, e.g., in a fixed follow-up setting, this could be the most severe event for each patient). The estimate is a marginal estimate, meaning that it reflects a win odds across all pairs of patients, rather than for a pair with identical covariate values. In Appendix B, we extend this approach to calculate the adjusted win ratio. Here a comparable adjustment to that in (2) is carried out, but now separately for the adjusted win probability and loss probability. We note that when using this method for estimating the adjusted win ratio, a single estimate of the association between covariate and outcome cannot be obtained, unlike when estimating the adjusted win odds. In contrast, when using the ordinal logistic regression-based approach for estimating the adjusted win ratio, quantifying the prognostic value of covariates is straightforward.

### 4.3. Inverse probability weighting (IPW)

Wang *et al*. ^13^ describe a covariate adjustment approach based on inverse probability weighting (IPW) using propensity scores. A propensity score is first calculated for each patient representing the probability of being assigned to the intervention group given their baseline characteristic, e.g. using logistic regression ^24^.

It may seem counter-intuitive to model treatment assignment in a randomized trial, where the theoretical probability of assignment is known and fixed by the randomization scheme (e.g., 0.5 for 1:1 randomization). However, using covariates to predict these probabilities within the trial sample, and weighting patients based on these probabilities, will take into account any chance imbalances in baseline covariates. This approach has been shown to reduce variance in trials using conventional outcomes ^25^.

Once the propensity scores are estimated, they are used as weights in the analysis. Wang et al. explore alternative weighting schemes: here we focus on one specific implementation. Let *P*_*i*_ be the propensity score for patient *i* in the intervention group and *P*_*j*_ be the propensity score of patient *j* in the control group, then the weight assigned to the comparison of patient *i* to patient *j* is 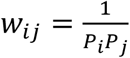. The win ratio is then calculated as the weighted sum of wins divided by the weighted sum of losses, i.e. 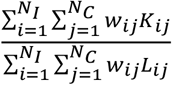. In this equation,

- *N*_*I*_ and *N*_*C*_ are the number of patients in the intervention group and control group, respectively.
- *K*_*ij*_ = 1 if patient *i* (intervention) wins against patient *j* (control), and 0 otherwise
- *L*_*ij*_ = 1 if patient *i* loses against patient *j*, and 0 otherwise

The win difference and win odds can be calculated using an analogous approach. The underlying principle is that the weighted groups are balanced with respect to baseline characteristics. The method is a marginal estimator, so that the interpretation of win statistics remains unchanged through covariate adjustment.

### 4.4. Matching

Another straightforward approach to covariate adjustment is matching, as highlighted in the original win ratio article ^2^, where each patient from the intervention arm is paired with a similar patient from the control arm (assuming 1:1 randomization). This approach has proved unpopular because: i) it is difficult to pre-specify in advance which patients will be matched across arms in a transparent and reproducible way; (ii) there is usually slightly unequal patient numbers in the two arms, meaning a small number of patients will be unmatched and excluded from the analysis ^26^. Matching also performed poorly in preliminary simulation investigations, and we therefore do not pursue this method further.

## 5. Application in EMPEROR-Preserved

The EMPEROR-Preserved trial ^14^ randomised 5988 patients having heart failure with preserved ejection fraction to empagliflozin or placebo. The primary composite endpoint was time to first heart failure hospitalisation (HFH) or cardiovascular (CV) death, and patients were followed up for a median of 26 months. In addition to the primary trial results, a subsequent publication created the EMPEROR-Preserved risk score ^27^, a multivariable model which predicts individual patient risk of the primary outcome.

In this paper we compare results with and without covariate adjustment (using our ordinal logistic regression-based approach) when using a hierarchical composite outcome analogous to the primary outcome with a hierarchy of (1) cardiovascular death (2) HFH. We calculate results using an (i) unadjusted win ratio, (ii) an analysis adjusting only for log NT-proBNP, the strongest single predictor the EMPEROR-Preserved risk score; (iii) all available covariates included in the EMPEROR-Preserved risk score ^27^. For comparison with conventional covariate adjustment, we obtain analogous results using a Cox proportional hazards model for the primary outcome of time to first of CV death or HFH. Note that in both analyses, patients determined to have died from non-CV causes are treated as censored. Results are shown in Table 1.

**Table 1:**
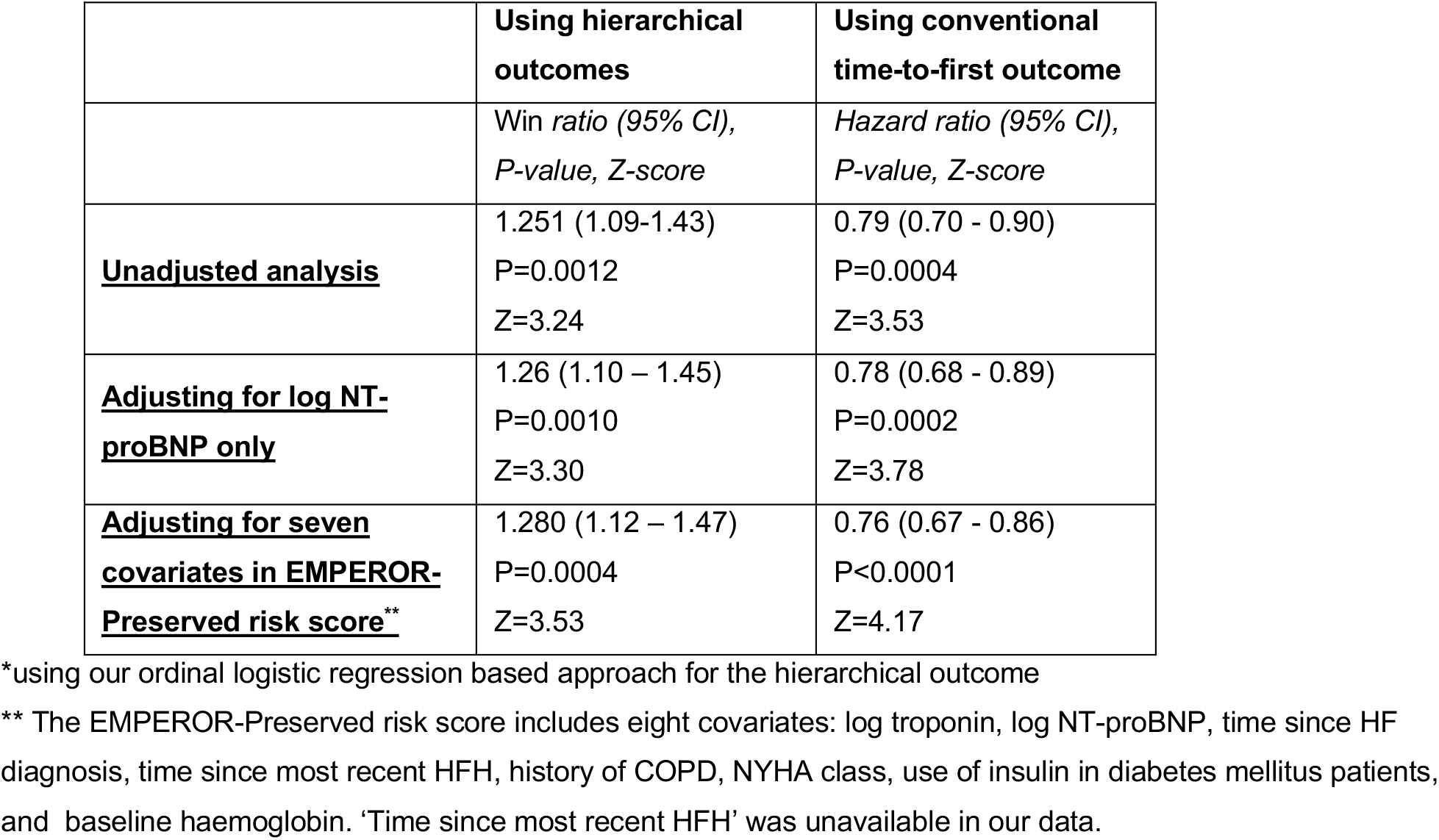
Results from EMPEROR-Preserved with and without covariate adjustment* for a hierarchical outcome of 1) time to cardiovascular death and 2) time to first HFH, and Cox model for composite time to cardiovascular death or HFH.

The unadjusted win ratio was 1.251 (95% CI, 1.09-1.43: p=0.0012 z=3.24). The win ratio moved further from the null value of 1.0 and statistical significance was enhanced upon adjustment, first for log NT-proBNP (win ratio=1.260 [95% CI 1.10 – 1.45, z=3.30] and then for all individual risk factors included in the EMPEROR-Preserved risk score (win ratio=1.280, 95% CI=1.12-1.47, z=3.53). A similar pattern, of estimates further from the null and increased statistical significance, was seen when using a Cox proportional hazards model for the conventional time-to-first event primary outcome (Table 1). Both the increase in statistical significance and change in point estimate are expected; adjusting for a prognostic covariate improves statistical power, and both the win ratio and Cox hazard ratio are non-collapsible, so the covariate-adjusted estimates will generally be larger in magnitude (further from the null) compared to the unadjusted estimates ^10,28^.

A useful feature of the covariate adjusted approach is that it also provides estimates for each covariate, indicating the strength of relationship with the primary outcome. With our ordinal model, these estimates are odds ratios under the proportional odds assumption. For example, in Supplementary Table 1, the coefficient for log NT-proBNP is 1.47: this means that a 1-unit increase in between-pair difference in log NT-proBNP increases the odds of having a loss (better outcome for control patient) by a factor 1.47 compared to the odds of having the same or better outcome (a tie or a win).

These results illustrate how our method works and shows its potential value. We further evaluate its properties in the simulations described below. More generally, note that the effect of covariate adjustment in any specific trial depends on strength of the covariate’s prognostic value and whether it is imbalanced between treatment groups.

## 6. Simulation study comparing methods for covariate adjustment

To further assess the effect of covariate adjustment, we ran simulations based on common hierarchical outcome structures in recent cardiovascular trials: (1) composite time-to-event outcomes and (2) composites mixing time-to-event and quantitative components. In each scenario, we simulated a normally distributed quantitative baseline covariate that was prognostic for the outcome, and also considered supplementary scenarios where the covariate was non-prognostic.

For each scenario we used 10,000 replications. We estimated the win ratio using five methods: unadjusted and using four covariate-adjusted approaches (including our proposed ordinal model). For each scenario, we reported the mean log win ratio (or log win odds for methods that cannot calculate the win ratio), empirical SE (Monte Carlo SD), median Z-statistic, median win ratio with 95% CI, statistical power (P < 0.05), and the estimated increase in effective sample size (calculated per ^9^, see Appendix C).

### 6.1. Simulations for a hierarchical composite based on time-to-event variable

#### Methods

We simulated a composite time-to-event outcome consisting of death and hospitalization. Simulations were loosely based on event rates from the ATTRIBUTE-CF trial ^29^, a trial in 632 patients with transthyretin cardiomyopathy, which used the win ratio for its primary analysis. While ATTRIBUTE-CF randomized patients 2:1 ratio to receive acoramidis hydrochloride or placebo, we used 1:1 randomization for simplicity. The treatment indicator was therefore generated from a Bernoulli distribution with probability 0.5. Time to death was simulated using an exponential baseline hazard. Parameters were set so that the control group had an approximately 40% death rate by 2.5 years (the trial follow-up period) and a hazard ratio of 0.7 in favour of the treatment group. Uniform administrative censoring was applied at 2.5 years (similar to the median duration of follow-up in ATTRIBUTE-CF). For time-to-first hospitalisation, the control group had an approximately 60% event rate and the treatment hazard ratio was 0.65. For simplicity, death and hospitalization times were simulated independently, with hospitalization times censored at either the time of death or at 2.5 years, whichever came first (i.e. if the simulated death time came before the hospitalization time, then this patient would be recorded to have an event for death, but not hospitalisation). We also simulated a normally distributed baseline covariate. A 1-SD increase in the baseline covariate was associated with a hazard ratio of 1.75 for both death and hospitalizations, consistent with published associations of log NT-proBNP and the composite of cardiovascular death or heart failure hospitalization in heart failure trials ^27^. We additionally simulated data with the covariate unrelated to both components of the primary outcome. To confirm type-I error control we also simulated data with no effect of treatment on either component of the hierarchical outcome. Each simulated trial included 510 patients, a sample size chosen to achieve approximately 85% power using the unmatched win ratio. In each replicate we analyzed data using the win ratio with the outcome hierarchy defined as 1) time to all-cause death 2) time to first CV hospitalisation. To benchmark the effect of covariate adjustment in hierarchical outcomes settings, we also analysed each dataset using a Cox proportional hazards model with and without covariate adjustment. Further characteristics including the parameters used for simulation are given in Supplementary Table 2, scenario 1.

#### Results

Results for a hierarchical composite of two time-to-event components are shown in Table 2 and Supplementary Tables 3, 4, 5 and 6. In the central scenario, where the covariate was prognostic for the primary outcome with a hazard ratio of 1.75 per 1-SD increase, there was a considerable benefit in statistical power from covariate adjustment (Table 2). Using the unadjusted win ratio yielded a median win ratio of 1.457, a median Z-statistic of 3.02 and a statically significant result in 86.1% of simulated trials. Upon covariate adjustment, the median Z-statistic increased similarly irrespective of which method of covariate adjustment was applied to 3.23-3.26 and the statistical power to 89.7%-90.3%, representing an approximate 4% gain in power, equal to the increase in power resulting from an approximately 15% larger sample size. For the ordinal logistic approach and probability index models, which target conditional estimands, the median estimated win ratio (or win odds) was further away from the null value of 1 than the median unadjusted estimate. The median win ratio was 1.524 using our ordinal logistic regression approach, and the average win odds from a probability index model was 1.381 (versus 1.344 without adjustment: Supplementary Table 3). The movement further away from the null value is analogous to what happens when conventional covariate adjustment methods are used for binary or time-to-event outcomes. However, for the IPW and randomization-based methods, which estimate a marginal win ratio (or win odds), the median estimated win ratio was the same as the median unadjusted estimate (1.457), with the gain in power instead resulting from a reduction in the standard error.

**Table 2:** Statistical power for win ratio analyses with and without covariate adjustment for a prognostic covariate (HR=1.75 per 1-SD) in an hierarchical composite of death and hospitalization.

| *Data characteristics* |  |  |  |  |  |
| --- | --- | --- | --- | --- | --- |
|  | **N** | **% deaths intervention** | **% death controls** | **% hospitalisations intervention** | **% hospitalisations control** |
|  | 510 | 30.5% | 39.9% | 47.7% | 60.4% |
| *Simulations results* |  |  |  |  |  |
|  | **Mean log win ratio (SE)** | **Median Z-statistic** | **Median estimated win ratio and 95% confidence limits** | **Power, %** | **Effective increase in sample size** |
| Unadjusted | 0.378<br>(0.125) | 3.023 | 1.457 (1.142 to 1.861) | 86.1% | Reference |
| **Adjusted for prognostic covariate** |  |  |  |  |  |
| Ordinal | 0.422<br>(0.129) | 3.255 | 1.524 (1.182 to 1.963) | 90.3% | 15.9% |
| Randomisation-based | 0.378<br>(0.117) | 3.226 | 1.457 (1.159 to 1.831) | 89.7% | 13.9% |
| IPW | 0.378<br>(0.116) | 3.243 | 1.457 (1.16 to 1.83) | 89.8% | 14.2% |

To benchmark the benefits of covariate adjustment when using the win ratio to those when using conventional outcomes, we analysed the data using a time-to-first event approach and the Cox proportional hazards model (Supplementary Table 4). When using Cox models, the statistical power increased from 87.4% in unadjusted models to 89.9% in adjusted models, equivalent to an increase in sample size of 7.0%. The benefits of covariate adjustment with the win ratio therefore appear similar or greater than the benefit when using conventional time-to-event outcomes. In Supplementary Table 5 we show results when using the win ratio with and without adjustment for a covariate that is not prognostic (i.e. unrelated to the outcome). Power was similar to when using an unadjusted approach, illustrating that there is no meaningful loss in statistical power from using covariate adjustment in this context. In Supplementary Table 6 we present results for the win ratio when adjusting for a prognostic covariate in the absence of a treatment effect, showing that for all methods, including our ordinal regression-based method, the type-1 error rate is maintained under the null hypothesis.

### 6.2. Hierarchical composite outcome including a quantitative component

#### Methods

To explore this scenario, we simulated data loosely based on the EMPULSE trial ^30^, which evaluated the effect of empagliflozin in 510 patients with acute heart failure after 90 days of treatment. The win ratio analysis in EMPULSE used three main components: 1) death from any cause; 2) heart failure events (HFE); and 3) Kansas City Cardiomyopathy Questionnaire (KCCQ) Overall Symptom score; a quantitative measure of quality of life that has been widely validated in heart-failure trials.

In our simulations, time to all-cause death in the control group followed an exponential distribution with parameters chosen so that ~5% of patients died before administrative censoring at 0.25 years. The treatment group was assigned a hazard ratio of 0.8. Time to first heart failure event was also simulated using an exponential distribution, independent of time to all-cause death, targeting a ~17% event rate in the control group, with a hazard ratio of 0.7 for intervention. Baseline and follow-up KCCQ scores (measured at 90 days) were simulated from a bivariate normal distribution bounded between 0 and 100. The baseline KCCQ score had a mean of 40. At follow-up, the mean score was 74 in the intervention and 70 in the control group, with a common standard deviation of 17. Since the gain in statistical power from covariate adjustment depends on the correlation between baseline and follow-up, we considered three levels of correlations between the KCCQ measurements: of 0.25, 0.5, and 0.75. In all scenarios, the KCCQ scores were simulated to be independent of time to death or hospitalization, which – while not realistic –allowed us to isolate the specific contribution of adjusting for a baseline measure of a quantitative component outcome. We simulated 10,000 trials, with 460 patients per trial and 1:1 allocation, a sample size chosen to yielded approximately 85% power. Key simulation characteristics are summarised in Supplementary Table 2, scenario 2.

In each simulated database we estimated the win ratio using an unmatched approach and covariate adjustment methods using a hierarchy of 1) time to death; 2) time to hospitalization; and 3) KCCQ score at follow-up. We also conducted additional unmatched analyses but with a modified third level of the hierarchy of: (i) change from baseline in KCCQ score: as used in the EMPULSE trial ^30^ and TRILUMINATE trial ^31^; (ii) the residual of a linear regression of KCCQ score at follow-up on KCCQ score at baseline. These latter implementations alter the estimand and the interpretation of the win ratio, a matter we return to later.

#### Results

Table 3 shows the simulation results for this 3-level hierarchical composite. We note with this hierarchical outcome, the majority of decisions are made on the basis of KCCQ, which is the quantitative outcome at the bottom of the hierarchy. The gain in statistical power from using covariate adjustment was dependent on the strength of relationship between the baseline KCCQ and end of study KCCQ. Regardless of the choice of method for covariate adjustment there was approximately: (i) a 5-6% increase in effective sample size when the correlation between baseline and end of study measurements was 0.25; (ii) a 33% increase in effective sample size when the correlation was 0.5; (ii) an 80-90% increase in effective sample size when the correlation was 0.75.

**Table 3:** Statistical power for win ratio analyses with and without adjustment for baseline KCCQ, across varying baseline-follow-up correlations strengths (weak=0.25, moderate=0.5, strong=0.75).

**a) Data characteristics**
|  | N | % death<br>s<br>interv<br>ention | % deaths<br>control<br>s | % hospitali<br>sations<br>intervent<br>ion | % hospital<br>isations<br>control | KCCQ at<br>follow-up<br>mean (SD)<br>interventio<br>n | KCCQ at<br>follow-up<br>mean (SD)<br>control |
| --- | --- | --- | --- | --- | --- | --- | --- |
| Weak correlation (0.25) | 500 | 5.82% | 4.70% | 12.4% | 17.3% | 73.2 (16.1) | 69.0 (17.1) |
| Weak correlation (0.5) | 500 | 5.82% | 4.69% | 12.4% | 17.3% | 73.4 (15.9) | 69.2 (16.9) |
| Strong correlation (0.75) | 500 | 5.80% | 4.71% | 12.4% | 17.2% | 73.6 (15.5) | 69.4 (16.6) |

**b) Simulation results**
|  | Mean log<br>win ratio<br>(SE) | Median<br>Z-statistic | Median estimated<br>win ratio and 95%<br>confidence limits | Power,<br>% | Effective<br>increase in<br>sample size |
| --- | --- | --- | --- | --- | --- |
| **Weak correlation** |  |  |  |  |  |
| Unadjusted | 0.306 (0.106) | 2.892 | 1.359 (1.104 to 1.673) | 82.3% | Reference |
| Unadjusted using<br>change from baseline | 0.274 (0.105) | 2.613 | 1.315 (1.071 to 1.614) | 74.6% | -18.4% |
| Unadjusted using<br>residuals | 0.317 (0.106) | 3.001 | 1.375 (1.117 to 1.692) | 84.8% | 7.7% |
| **Adjusted for KCCQ** |  |  |  |  |  |
| Ordinal | 0.318 (0.107) | 2.973 | 1.374 (1.114 to 1.695) | 84.6% | 5.7% |
| Randomisation-based | 0.311 (0.105) | 2.972 | 1.365 (1.112 to 1.676) | 84.4% | 5.6% |
| IPW (Duolao) | 0.311 (0.105) | 2.972 | 1.365 (1.112 to 1.676) | 84.4% | 5.6% |
| Probability index model* | 0.318 (0.107) | 2.97 | 1.374 (1.114 to 1.695) | 84.5% | 5.5% |
| **Moderate correlation** |  |  |  |  |  |
| Unadjusted | 0.308 (0.104) | 2.97 | 1.361 (1.111 to 1.668) | 84.4% | Reference |
| Unadjusted using<br>change from baseline | 0.351 (0.104) | 3.359 | 1.419 (1.157 to 1.74) | 92.2% | 27.9% |
| Unadjusted using<br>residuals | 0.37 (0.104) | 3.557 | 1.448 (1.181 to 1.775) | 94.7% | 43.4% |
| **Adjusted for KCCQ** |  |  |  |  |  |
| Ordinal | 0.366 (0.107) | 3.422 | 1.443 (1.169 to 1.779) | 93.0% | 32.8% |
| Randomisation-based | 0.333 (0.097) | 3.435 | 1.395 (1.154 to 1.686) | 93.2% | 33.7% |
| IPW (Duolao) | 0.333 (0.097) | 3.435 | 1.395 (1.153 to 1.686) | 93.2% | 33.7% |
| Probability index model* | 0.366 (0.107) | 3.422 | 1.442 (1.169 to 1.779) | 93.0% | 32.7% |
| **Strong correlation** |  |  |  |  |  |
| Unadjusted | 0.309 (0.104) | 2.959 | 1.362 (1.11 to 1.671) | 84.0% | Reference |
| Unadjusted using<br>change from baseline | 0.512 (0.107) | 4.769 | 1.666 (1.351 to 2.055) | 99.8% | 159.6% |
| Unadjusted using<br>residuals | 0.517 (0.107) | 4.828 | 1.675 (1.359 to 2.066) | 99.8% | 166.2% |
| **Adjusted for KCCQ** |  |  |  |  |  |
| Ordinal | 0.461 (0.113) | 4.069 | 1.584 (1.269 to 1.976) | 98.5% | 89.0% |
| Randomisation-based | 0.366 (0.088) | 4.175 | 1.443 (1.215 to 1.714) | 98.7% | 99.0% |
| IPW (Duolao) | 0.366 (0.088) | 4.17 | 1.442 (1.214 to 1.712) | 98.7% | 98.6% |
| Probability index model* | 0.461 (0.113) | 4.069 | 1.584 (1.269 to 1.976) | 98.5% | 89.0% |

\* the probability index model estimates the adjusted win odds. In this scenario, in the absence of ties, this reduces to the win ratio.

Using change from baseline KCCQ had mixed fortunes, resulting in a loss of power when the correlation between KCCQ at baseline and at follow-up was weak (18% decrease in effective sample size), a gain when the correlation was moderate (28% increase) and a substantial gain when the correlation was strong (160% increase). Using the residuals from a linear regression of KCCQ at follow-up on KCCQ at baseline yielded increases in effective sample size of 8%, 43% and 166% for a weak, moderate, and strong correlation, respectively, and was therefore more efficient than using covariate adjustment. In the artificial scenario where KCCQ is the only outcome component in the hierarchy (Supplementary Table 7), the gain in power when adjusting for KCCQ at baseline as a covariate and when using change of residuals was very similar.

## Discussion

Covariate adjustment is widely used in randomized controlled trials to improve statistical power and precision ^9,10,32^ and to target conditional effects. Regulatory guidance from agencies such as the Food and Drug Administration ^33^ and the European Medicines Agency ^34^ also supports its use, though the FDA notes that in non-collapsible models, adjustment address a different estimand: instead of a population-average to a conditional effect. While well established for conventional time-to-first composite outcomes, covariate adjustment has been less explored for hierarchical composite outcomes, such as those analyzed with the win ratio. There is a growing need for these methods since the win ratio is becoming a common primary outcome in randomized trials, especially in cardiovascular disease ^5^. A lack of a clear illustration of the benefits of covariate adjustment in the context of hierarchical outcomes and a lack of software implementations that accommodate such multi-level outcomes is a missing piece preventing the broader adoption and routine use of the win ratio in randomized trial analysis, particularly in settings where adjustment offers meaningful gains in precision.

While several methods exist for covariate adjustment with hierarchical outcomes, they do not fully address key analytical needs. Probability index models can quantify the relationship between a covariate and the outcome, but are restricted to estimating the win odds. In contrast, the IPW and randomization-based approaches can be used to estimate the win ratio, but they do not readily give insight into the specific relationship between a covariate and the hierarchical outcome. For the randomization-based approach, while the original method for the win odds allows for such a quantification, obtaining a similar single estimate for the win ratio is not straightforward. Consequently, a methodological gap exists for a method that can both estimate the win ratio and quantify the influence of covariates on the outcome.

To address this gap, we developed a new covariate adjustment method for hierarchical outcomes using ordinal regression. We compared it with probability index models, inverse probability weighting (IPW), and randomization-based methods—in scenarios commonly seen in clinical trials. These included adjustment for a strong prognostic covariate and for a baseline measure of a component of the outcome. Our simulations showed that adjusting for a prognostic baseline variable or outcome component consistently improves power, while adjusting for a non-prognostic covariate does not harm the analysis. Overall, all four adjusted win ratio estimators produced power gains similar to those observed with covariate adjustment for conventional outcomes. These findings suggest that hierarchical outcomes behave similarly to other types when covariate adjustment is applied and that the benefits are clearest when covariates are prognostic. While our scenarios were limited, they reflect common trial settings and are likely to generalize.

Among the methods we evaluated, our proposed ordinal regression approach offers distinct advantages. Like a Cox model, it provides a conditional estimate of the treatment effect, accounting for individual covariates. It also allows estimation of the prognostic impact of each covariate on the hierarchical outcome. A limitation of the ordinal adjusted win ratio is that, like a conventional Cox model or a logistic regression, it is not robust to model misspecification, as it estimates a conditional effect measure, though the type-1 error rate is maintained. Specifically, the proportional odds assumption will be violated when there is effect modification. A conditional estimate can also be obtained from the probability index model but this estimates a win odds and not a win ratio, limiting its utility. In contrast, the IPW and randomization-based methods estimate marginal effects that reflect average treatment effects in the population. The choice between conditional and marginal estimands is an ongoing debate. Supporters of conditional estimands argue they are more informative for clinical decisions, as they are more tailored to patient characteristics ^35^, while proponents of marginal effects argue they may be preferable because they can be defined without reference to specific modelling assumptions, and can be estimated unbiasedly even if models misspecify the effects of covariates ^36,37^. Ultimately, it is a trial’s objectives that should guide the choice of an estimand.

In our paper, we develop a new methodology for estimating a conditional covariate-adjusted win ratio using an ordinal approach, and extend the marginal randomisation-based method for adjustment, which was previously only defined for the win odds, to the win ratio. The win ratio is a widely used measure of relative treatment benefit, with a clear clinical interpretation. For instance, a win ratio of 1.5 simply means that amongst untied pairs, the intervention treatment patient has a favourable outcome 1.5 times more often than the control patient. While the concern may be raised that the win ratio, unlike the win odds, ignores ties—leading to the same win ratio being estimated when e.g. 90% of patients are tied or when only 10% are tied—this issue is overcome by reporting the win difference alongside the win ratio, for a complementary absolute measure of effect, in the same way one commonly reports a risk difference alongside a risk ratio ^3^. Additionally note that while the win ratio point estimate does not explicitly incorporate ties, these are reflected in the confidence interval; a higher proportion of ties reduces the effective sample size, which increases the SE and widens the confidence interval ^38^.

Alternative approaches to covariate-adjustment, such as adjusting for the baseline quantitative outcome using change scores or residuals, are possible. Using the residuals was more efficient than covariate adjustment. However, we note that this may be a feature specific to our simulation setup, whereby the baseline value of the quantitative outcome was only associated with the follow-up value and was independent of other outcomes in the hierarchy. Whether this is realistic in practice would depend on the specific implementation. In the covariate adjustment approach, a single adjustment is performed across all levels of the hierarchy. Because the higher-priority components are independent of baseline KCCQ, this results in less overall power gain than when substituting follow-up KCCQ in the hierarchical outcome with KCCQ residuals from a linear regression. This is supported by our finding that when the hierarchy consists only of the quantitative KCCQ component, the regression-based and residual-based adjustments result in nearly identical power gains. A drawback of the residual-based approach is that it addresses a different estimand and complicates interpretation of the win ratio. Specifically, the definition of a ‘win’ shifts from having a better KCCQ score at follow-up to having a greater improvement from baseline KCCQ so that, for a given pair, the treatment group patient could be classified as a ‘win’ despite having a lower absolute score at follow-up than the placebo patient. In contrast, the covariate-adjustment methods maintain the outcome definition.

Although our focus was on randomized trials, these methods are also relevant to observational analyses, where covariate adjustment is used primarily to adjust for confounding rather than to improve precision. Currently, software options for covariate-adjusted win ratio methods are limited. The R packages *hce* ^23^ and *sanon* ^22^ implement the randomisation-based method for adjusted Mann–Whitney probabilities, and the *pim* package ^19^ implements the probability index model approach. While these packages can be used to obtain adjusted win odds, they do not compute adjusted win ratios and are limited to univariate HCEs (multiple outcomes that can be collapsed into a single distribution). We plan to update the *winratiotest* command in Stata ^39^ to support our ordinal adjustment method. In the meantime, we provide worked examples in R ^40^ for both our ordinal method and the randomization-based method in the Supplementary Materials to support implementation.

In conclusion, covariate adjustment is just as beneficial for hierarchical composite outcomes as for more traditional ones, when covariates are prognostic. Our ordinal regression method for the win ratio is straightforward to apply, performs as well as existing approaches, and, unlike these, additionally quantifies the prognostic impact of covariates on the hierarchical outcome. With greater accessibility through worked examples and future software updates, this method can help bring the benefits of covariate adjustment to a wider range of trials using hierarchical outcomes.

## Data Availability

The data used in this study were provided by Boehringer Ingelheim and accessed via the Vivli Inc. platform. The authors are not permitted to share the data under the terms of the Data Use Agreement. De-identified individual participant data and clinical study documents from the EMPEROR-Preserved trial are available to researchers upon approval of a research proposal submitted through the Vivli portal.

## Acknowledgements

This publication is based on research done under AstraZeneca post-doctoral funding. This publication is based on research using EMPEROR-Preserved trial data from data contributors Boehringer Ingelheim that has been made available through Vivli, Inc. Vivli has not contributed to or approved, and is not in any way responsible for, the contents of this publication.

