## Supplement for "Covariate adjustment for hierarchical outcomes and the win ratio: how to do it and is it worthwhile?"

*Supplementary Tables* 2

Supplementary table 1. Results in the EMPEROR-Preserved trial for the ordinal adjusted win ratio 2

Supplementary Table 2: Overview of key characteristics of simulation scenarios 3

Supplementary Table 3: Statistical power for win odds analyses with and without covariate adjustment for a prognostic covariate 4

Supplementary Table 4: Statistical power for Cox proportional hazards analyses with and without covariate adjustment for a prognostic covariate 4

Supplementary Table 5: Statistical power for win ratio analyses with and without covariate adjustment for a non-prognostic covariate 5

Supplementary Table 6: Type-1 error for win ratio analyses without treatment effect (under the null hypothesis), with and without covariate adjustment for a prognostic covariate 6

Supplementary table 7: Statistical power for win ratio analyses for a single quantitative outcome 7

*Appendix A: Derivation of a standard error for the adjusted ordinal win ratio, estimator properties in simulation and a worked example.* 8

A.1 Derivation of a standard error for the adjusted win ratio 8

A.2 Estimator properties in simulation 9

A.3 A worked example 12

*Appendix B: Adaptation of the randomisation-based method to calculate the adjusted win ratio and win difference* 18

B.1 Deriving an SE for the adjusted win ratio 20

B.2 Estimator properties in simulation 21

B.3 Adjusting for multiple covariates 23

B.4. Worked example 24

*Appendix C: Calculation of effective increase in sample size based on Z-statistic from simulations* 30

**Supplementary table 1**. Results in the EMPEROR-Preserved trial for the ordinal adjusted win ratio (hierarchical outcome of 1) time to CV death 2) time to first HFH) and Cox proportional hazards regression (time to first of CV death or HFH): covariate estimates.

| | **Using hierarchical outcomes (ordinal win ratio)**<br>*Odds ratio* (95% CI), P-value, Z-score* | **Using conventional time-to-first outcome (Cox)**<br>*Hazard ratio (95% CI), P-value, Z-score* |
|---|---|---|
| *Adjusting for log NT-proBNP alone* | | |
| **Log NT-proBNP (pg/ml)** | 1.47(1.41-1.54)<br>P<0.0001<br>Z=17.56 | 1.93 (1.79 - 2.07)<br>P<0.0001<br>Z=17.92 |
| *Adjusting for all covariates in the EMPEROR-Preserved risk score* | | |
| **Log NT-proBNP (pg/ml)** | 1.34 (1.28-1.41)<br>P<0.0001<br>Z=12.71 | 1.64 (1.52 – 1.77)<br>P<0.0001<br>Z=12.69 |
| **Log troponin (ng/L)** | 1.40 (1.31-1.5)<br>P<0.0001<br>Z=9.51 | 1.60 (1.46 – 1.76)<br>P<0.0001<br>Z=9.76 |
| **NYHA class III/IV** | 1.48 (1.31-1.68)<br>P<0.0001<br>Z=6.32 | 1.71 (1.48 – 1.97)<br>P<0.0001<br>Z=7.44 |
| **History of COPD** | 1.47 (1.27-1.69)<br>P<0.0001<br>Z=5.32 | 1.73 (1.48 – 2.03)<br>P<0.0001<br>Z=6.80 |
| **Use of insulin in DM patients** | | |
| Non-DM | 1.00 (reference) | |
| DM + insulin | 1.39 (1.21-1.60)<br>P<0.0001<br>Z=4.74 | 1.69 (1.42 – 2.01)<br>P<0.0001<br>Z=5.92 |
| DM + no insolin | 1.06 (0.98-1.15)<br>P=0.16<br>Z=1.41 | 1.16 (1.00 – 1.34)<br>P=0.052<br>Z=1.95 |
| **Baseline haemoglobin** | | |
| ≥ 12 g/dl | 1.00 (reference) | |
| < 12 g/dl | 1.31 (1.17-1.47)<br>P<0.0001<br>Z=4.6 | 1.47 (1.27 – 1.70)<br>P<0.0001<br>Z=5.22 |
| **Time since HF diagnosis** | 1.00 (reference) | |
| 3 months-1 year | 1.11 (1.02-1.21)<br>P=0.011<br>Z=2.53 | 1.26 (1.07 – 1.47)<br>P=0.0045<br>Z=2.84 |

*Odds ratios under the proportional odds assumption, comparing the odds of having a loss to the odds of having a tie or a win, for a one-unit increase in within-pair covariate difference

**Supplementary Table 2:** Overview of key characteristics of simulation scenarios

| | | | | Death | | | Hospitalisation | | | KCCQ at follow-up | | | |
|---|---|---|---|---|---|---|---|---|---|---|---|---|---|
| | | | | | Average % with death | | | Average % with hospitalisation | | | | KCCQ means (SD) at follow-up after bounding** | |
| **Scenario** | **Composite outcome** | **Number of patients per simulated trial** | **Follow-up** | Rate parameter used in simulation* | Intervention | Control | Rate parameter used in simulation* | Intervention | Control | Bivariate normal distribution** for KCCQ at baseline ($KCCQ_b$) and KCCQ at follow-up ($KCCQ_b$) | mean cor($KCCQ_b$, $KCCQ_f$) after bounding** | Intervention | Control |
| 1: prognostic covariate | Death, hospitalization | 510 | 2.5 year | 0.19 *exp( log(0.7)*Ti + log(1.75)*$X_i$) | 30.5% | 39.9% | 0.37 *exp( log(0.7)*Ti + log(1.75)*$X_i$) | 47.7% | 60.4% | | | | |
| 1: not prognostic covariate | Death, hospitalization | 510 | 2.5 year | 0.19 *exp( log(0.7)*Ti) | 29.5% | 39.4% | 0.37 *exp( log(0.7)*$T_i$) | 46.0% | 61.3% | | | | |
| 2: weak correlation | Death, hopsitalization, KCCQ | 460 | 3 months | 0.24 *exp( log(0.7)*Ti) | 5.84% | 4.68% | 0.76 *exp( log(0.8)*Ti) | 17.3% | 12.4% | $\begin{pmatrix} KCCQ_b \\ KCCQ_f \end{pmatrix} \sim N\left[\begin{pmatrix} 40 \\ 72+6*T \end{pmatrix}, \begin{pmatrix} 400 & 128 \\ 128 & 400 \end{pmatrix}\right]$ | 0.25 | 73.2 (16.1) | 69.0 (17.1) |
| 2: moderate correlation | Death, hopsitalization, KCCQ | 460 | 3 months | 0.24 *exp( log(0.7)*Ti) | 5.82% | 4.69% | 0.76 *exp( log(0.8)*Ti) | 17.3% | 12.4% | $\begin{pmatrix} KCCQ_b \\ KCCQ_f \end{pmatrix} \sim N\left[\begin{pmatrix} 40 \\ 72+6*T \end{pmatrix}, \begin{pmatrix} 400 & 240 \\ 240 & 400 \end{pmatrix}\right]$ | 0.50 | 73.4 (15.6) | 69.2 (16.9) |
| 2: strong correlation | Death, hopsitalization, KCCQ | 460 | 3 months | 0.24 *exp( log(0.7)*Ti) | 5.82% | 4.70% | 0.76 *exp( log(0.8)*Ti) | 17.3% | 12.4% | $\begin{pmatrix} KCCQ_b \\ KCCQ_f \end{pmatrix} \sim N\left[\begin{pmatrix} 40 \\ 72+6*T \end{pmatrix}, \begin{pmatrix} 400 & 334 \\ 334 & 400 \end{pmatrix}\right]$ | 0.75 | 73.6 (15.5) | 69.4 (16.8) |

*With treatment, $T_i$ , taking the value 0 for the control group and 1 for the intervention group; and covariate $X_i$

**The values for $KCCQ_b$ and $KCCQ_f$ were bounded [0,100] after being drawn from bivariate normal distribution

**Supplementary Table 3**: Statistical power for win odds analyses with and without covariate adjustment for a prognostic covariate (HR=1.75 per 1-SD) for a hierarchical composite of death and hospitalization.

| | **Mean log win odds (SE)** | **Median Z-statistic** | **Median estimated win odds and 95% confidence limits** | **Power, %** | **Effective increase in sample size** |
|---|---|---|---|---|---|
| Unadjusted | 0.297 (0.098) | 3.023 | 1.344 (1.11 to 1.627) | 86.1% | Reference |
| **Adjusted for prognostic covariate** | | | | | |
| Ordinal | 0.323 (0.099) | 3.255 | 1.380 (1.137 to 1.674) | 90.3% | 15.9% |
| Randomisation-based | 0.297 (0.091) | 3.226 | 1.345 (1.124 to 1.608) | 89.7% | 13.9% |
| IPW | 0.296 (0.091) | 3.243 | 1.344 (1.124 to 1.607) | 89.8% | 14.2% |
| Probability index model | 0.323 (0.099) | 3.245 | 1.381 (1.136 to 1.678) | 89.9% | 15.2% |

**Supplementary Table 4:** Statistical power for Cox proportional hazards analyses with and without covariate adjustment for a prognostic covariate (HR=1.75 per 1-SD) for a composite outcome of time to death or hospitalization

| | Mean log coefficient (SE) | Median Z-statistic | Median estimated HR and 95% confidence limits | Power, % | Effective increase in sample size |
|---|---|---|---|---|---|
| Unadjusted | -0.369 (0.119) | -3.027 | 0.691 (0.547 to 0.872) | 87.4% | Reference |
| Adjusted | -0.384 (0.114) | -3.131 | 0.681 (0.544 to 0.853) | 89.9% | 7.0% |

**Supplementary Table 5:** Statistical power for win ratio analyses with and without covariate adjustment for a non-prognostic covariate (unrelated to a hierarchical composite outcome of time to death or hospitalization)

| *Data characteristics* | | | | | |
|---|---|---|---|---|---|
| | **N** | **% deaths intervention** | **% death controls** | **% hospitalisations intervention** | **% hospitalisations control** |
| | 510 | 29.5% | 39.4% | 46.0% | 61.3% |
| *Simulations results: win ratio* | | | | | |
| | **Mean log win ratio (SE)** | **Median Z-statistic** | **Median estimated win ratio and 95% confidence limits** | **Power, %** | **Effective increase in sample size** |
| Unadjusted | 0.415 (0.127) | 3.258 | 1.512 (1.179 to 1.938) | 90.5% | N Reference |
| **Adjusted for prognostic covariate** | | | | | |
| Ordinal | 0.415 (0.127) | 3.257 | 1.513 (1.179 to 1.941) | 90.4% | -0.1% |
| Randomisation-based | 0.415 (0.127) | 3.258 | 1.512 (1.179 to 1.939) | 90.5% | 0% |
| IPW | 0.413 (0.126) | 3.278 | 1.511 (1.18 to 1.935) | 90.6% | 0.2% |
| *Simulations results: win odds* | | | | | |
| | **Mean log win odds (SE)** | **Median Z-statistic** | **Median estimated win odds and 95% confidence limits** | **Power, %** | **Effective increase in sample size** |
| Unadjusted | 0.323 (0.098) | 3.278 | 1.381 (1.138 to 1.674) | 90.6% | Reference |
| **Adjusted for prognostic covariate** | | | | | |
| Probability index model | 0.323 (0.099) | 3.275 | 1.381 (1.138 to 1.675) | 90.7% | -0.2% |

**Supplementary Table 6:** Type-1 error for win ratio analyses without treatment effect (under the null hypothesis), with and without covariate adjustment for a prognostic covariate (HR=1.75 per 1-SD) in an hierarchical composite of death and hospitalization

| *Data characteristics* | | | | |
|---|---|---|---|---|
| **N** | **% deaths intervention** | **% death controls** | **% hospitalisations intervention** | **% hospitalisations control** |
| 510 | 39.8% | 39.9% | 60.4% | 60.4% |
| *Simulations results* | | | | |
| | **Mean log win ratio (SE)** | **Median Z-statistic** | **Median estimated win ratio and 95% confidence limits** | **Power/ Type-1 error rate %** |
| Unadjusted | 0.001 (0.117) | 0.019 | 1.002 (0.797 to 1.26) | 0.025 |
| Ordinal | 0.001 (0.121) | 0.006 | 1.001 (0.789 to 1.27) | 0.026 |
| Randomisation-based | 0.001 (0.109) | 0.009 | 1.001 (0.809 to 1.238) | 0.025 |
| IPW | 0.001 (0.108) | 0.007 | 1.001 (0.809 to 1.238) | 0.026 |

**Supplementary table 7:** Statistical power for win ratio analyses for a single quantitative outcome (KCCQ at follow-up), with and without baseline covariate adjustment, across varying baseline-follow-up correlations strengths (weak=0.25, moderate=0.5, strong=0.75).

| | Mean log win ratio (SE) | Median Z-statistic | Median estimated win ratio and 95% confidence limits | Power, % | Effective increase in sample size |
|---|---|---|---|---|---|
| **Weak correlation** | | | | | |
| Unadjusted | 0.287 (0.104) | 2.755 | 1.332 (1.086 to 1.634) | 78.7% | Reference |
| Unadjusted using change from baseline | 0.244 (0.104) | 2.338 | 1.277 (1.04 to 1.567) | 64.7% | -28.0% |
| Unadjusted using residuals | 0.303 (0.104) | 2.905 | 1.354 (1.104 to 1.661) | 82.7% | 11.2% |
| **Adjusted for KCCQ** | | | | | |
| Ordinal | 0.306 (0.106) | 2.895 | 1.358 (1.104 to 1.67) | 82.8% | 10.4% |
| Randomisation-based | 0.295 (0.101) | 2.897 | 1.342 (1.1 to 1.637) | 82.8% | 10.6% |
| IPW (Duolao) | 0.294 (0.101) | 2.898 | 1.342 (1.1 to 1.637) | 82.8% | 10.7% |
| Probability index model* | 0.306 (0.106) | 2.893 | 1.357 (1.104 to 1.67) | 82.7% | 10.3% |
| **Moderate correlation** | | | | | |
| Unadjusted | 0.29 (0.106) | 2.747 | 1.337 (1.087 to 1.645) | 77.7% | Reference |
| Unadjusted using change from baseline | 0.349 (0.104) | 3.326 | 1.416 (1.153 to 1.737) | 91.9% | 46.6% |
| Unadjusted using residuals | 0.376 (0.105) | 3.558 | 1.454 (1.183 to 1.787) | 94.9% | 67.8% |
| **Adjusted for KCCQ** | | | | | |
| Ordinal | 0.391 (0.11) | 3.528 | 1.474 (1.188 to 1.829) | 94.5% | 64.9% |
| Randomisation-based | 0.325 (0.091) | 3.552 | 1.382 (1.156 to 1.652) | 94.6% | 67.2% |
| IPW (Duolao) | 0.324 (0.091) | 3.55 | 1.381 (1.156 to 1.65) | 94.7% | 67.0% |
| Probability index model* | 0.391 (0.11) | 3.528 | 1.474 (1.188 to 1.829) | 94.5% | 64.9% |
| **Strong correlation** | | | | | |
| Unadjusted | 0.292 (0.105) | 2.779 | 1.341 (1.09 to 1.649) | 79.1% | Reference |
| Unadjusted using change from baseline | 0.573 (0.108) | 5.316 | 1.771 (1.435 to 2.186) | 100% | 266.0% |
| Unadjusted using residuals | 0.58 (0.108) | 5.371 | 1.782 (1.443 to 2.2) | 100% | 273.7% |
| **Adjusted for KCCQ** | | | | | |
| Ordinal | 0.627 (0.117) | 5.345 | 1.871 (1.487 to 2.354) | 100% | 270.0% |
| Randomisation-based | 0.372 (0.069) | 5.386 | 1.45 (1.266 to 1.659) | 100% | 275.7% |
| IPW (Duolao) | 0.37 (0.069) | 5.394 | 1.448 (1.266 to 1.657) | 100% | 276.9% |
| Probability index model* | 0.627 (0.117) | 5.347 | 1.871 (1.487 to 2.354) | 100% | 270.2% |

* the probability index model estimates the adjusted win odds. In this scenario, in the absence of ties, this reduces to the win ratio.

#### Appendix A: Derivation of a standard error for the adjusted ordinal win ratio, estimator properties in simulation and a worked example.

*In this appendix we derive an approximate standard error for the adjusted ordinal win ratio (Section A.1), illustrate the properties of the adjusted win ratio estimator (Section A.2), and provide a worked example for estimating the adjusted ordinal win ratio and its SE with R code (Section A.3).*

##### A.1 Derivation of a standard error for the adjusted win ratio

Let $f(\alpha_1, \alpha_0)$ denote the function of intercepts $\alpha_1$ and $\alpha_0$ which estimates the log adjusted win ratio:

$$f(\alpha_{0.5}, \alpha_0) = \log\left(\frac{1 - \frac{1}{1+\exp(-\alpha_1)}}{\frac{1}{1+\exp(-\alpha_0)}}\right)$$

where $\alpha_1$ and $\alpha_0$ are estimated from the ordinal logistic regression model, as described in the main text, with the adjusted win and loss probabilities estimated as $\hat{\lambda}_a = \frac{1}{1+\exp(-\alpha_0)}$ and $\hat{\vartheta}_a = 1 - \frac{1}{1+\exp(-\alpha_1)}$ , respectively.

$\text{SE}_{\log(\text{WR}_a)} = \sqrt{\text{var}[\log(\text{WR}_a)]}$ is estimated with the delta method, which uses the first order derivatives of $f(\alpha_1, \alpha_0)$ with respect to $\alpha_1$ and $\alpha_0$, and the covariance matrix of the untransformed intercept terms:

$$\text{var}[\log(\text{WR}_a)] \approx \begin{bmatrix}\frac{\partial f}{\partial \alpha_1} & \frac{\partial f}{\partial \alpha_0}\end{bmatrix} \times \Sigma \times \begin{bmatrix}\frac{\partial f}{\partial \alpha_1} & \frac{\partial f}{\partial \alpha_0}\end{bmatrix} =$$
$$\begin{bmatrix}\frac{-1}{\exp(\text{-}\alpha_1 + 1)} & \frac{-1}{\exp(\alpha_0) \times (1+\exp(-\alpha_0))}\end{bmatrix} \times \Sigma \times$$
$$\begin{bmatrix}\frac{-1}{\exp(\text{-}\alpha_1 + 1)} & \frac{-1}{\exp(\alpha_0) \times (1+\exp(-\alpha_0))}\end{bmatrix},$$

with

$$\Sigma = \begin{bmatrix} \sigma^2_{\alpha_1} & \sigma_{\alpha_1\alpha_0} \\ \sigma_{\alpha_1\alpha_0} & \sigma^2_{\alpha_0} \end{bmatrix}.$$

$\Sigma$ is the covariance matrix of the intercept terms from the ordinal logistic regression model, with $\sigma^2_{\alpha_1}$ the model-estimated variance of $\alpha_1$, $\sigma^2_{\alpha_0}$ the variance of $\sigma^2_{\alpha_0}$, and $\sigma_{\alpha_1\alpha_0}$ the covariance between $\alpha_1$ and $\sigma^2_{\alpha_0}$. $\Sigma$ can be estimated with standard statistical software, by employing a sandwich estimator that takes into account the two sources of clustering: each patient *i* in the treatment arm is repeated $N_I$ times, while each patient *j* in the control arm is repeated $N_C$ times. In Section A.3, we provide a worked example of estimating the ordinal adjusted win ratio with corresponding SE.

#### A.2 Estimator properties in simulation

We investigated the properties of the ordinal win ratio estimator in simulation, employing the same data-generating mechanism as in Section 4.1 of the main text, which describes a comparative simulation study for all estimators. In brief, we simulated data with a composite time-to-event outcome of death and hospitalization, loosely based on the rates of events observed in the ATTRIBUTE-CF trial, and a baseline covariate, log NT-proBNP, which is either prognostic (with a hazard ratio of 1.75 for both components of the outcome) or non-prognostic. We repeated the simulation in absence of a treatment effect (under the null).

For each scenario, we compared estimates for the unadjusted win ratio estimator to the ordinal win ratio estimator. For the latter, we considered both unadjusted estimates and covariate-adjusted estimates. Additionally, we compared the standard error, mean Z-statisic and power when using the Monte Carlo derived SE (i.e. standard deviation of log win ratio across all simulated datasets) and when the approximate SE is derived for each simulated dataset using the delta method as described in Section A.1. Results are shown in the table below.

**Appendix A Table:** Ordinal win ratio estimator properties in simulation, for a hierarchical composite outcome of time to death or hospitalization, with treatment effect and under the null, and with and without adjustment for a prognostic or non-prognostic covariate.

| | | | Using Monte Carlo SD | | | Using analytical SE | | |
|---|---|---|---|---|---|---|---|---|
| | Mean win ratio | Mean log win ratio | Log win ratio SE (MCSD) | Mean Z-statistic | Power | Mean win ratio SE (analytical) | Mean Z-statistic | Power/ type-1 error |
| *With treatment effect and prognostic covariate (HR=1.75 per 1 SD increase)* | | | | | | | | |
| **Unadjusted** | 1.471 | 0.378 | 0.125 | 3.033 | 0.861 | | | |
| **Ordinal, unadjusted** | 1.471 | 0.378 | 0.125 | 3.033 | 0.861 | 0.125 | 3.009 | 0.861 |
| **Ordinal, adjusted** | 1.538 | 0.422 | 0.129 | 3.263 | 0.902 | 0.130 | 3.245 | 0.905 |
| *With treatment effect and non- prognostic covariate* | | | | | | | | |
| **Unadjusted** | 1.526 | 0.415 | 0.127 | 3.27 | 0.905 | | | |
| **Ordinal, unadjusted** | 1.527 | 0.415 | 0.127 | 3.271 | 0.905 | 0.127 | 3.272 | 0.910 |
| **Ordinal, adjusted** | 1.527 | 0.415 | 0.127 | 3.268 | 0.904 | 0.127 | 3.270 | 0.909 |
| *With null treatment effect and prognostic covariate (HR=1.75 per 1 SD increase)* | | | | | | | | |
| **Unadjusted** | 1.008 | 0.001 | 0.117 | 0.012 | 0.025 | | | |
| **Ordinal, unadjusted** | 1.008 | 0.001 | 0.117 | 0.012 | 0.025 | 0.118 | 0.012 | 0.024 |
| **Ordinal, adjusted** | 1.009 | 0.001 | 0.121 | 0.010 | 0.026 | 0.122 | 0.010 | 0.026 |

Across all scenarios, the conventional unadjusted estimator and the unadjusted ordinal estimator behaved near identically. For the adjusted ordinal estimator, adjusting for a prognostic covariate resulted in an expected gain in power, while adjusting for a non-prognostic covariate gave very similar results to the unadjusted estimators. In the absence of a treatment effect, the one-sided type-1 error rate was 0.026 for the adjusted ordinal estimator. For all scenarios, the Monte-Carlo derived SE (standard deviation of log win ratio across all simulated datasets) was comparable to the mean of analytical SE computed for each simulated datasets. Using the analytical SE to calculate the Z-statistic and power gave very similar values to the ones obtained using the simulation SD. In sum, our simulation results indicate that the ordinal win ratio estimator is unbiased and controls the type-1 error.

#### A.3 A worked example

First, we simulate a sample dataset using the function fsim_data. For simplicity, we consider only a single time-to-event outcome with exponentially distributed survival times:

```
fsim_data <- function(seed, n){
 set.seed(seed)

  # ID indicator
ID <- seq(1,n)
  # treatment indicator
X <- rbinom(n = n, size = 1, prob = 0.5)
  # Covariate
C <- rnorm(n, mean=0.5, sd=1)

  # Generating xponentially distributed survival times, with base rate,
  # lambda, treatment effect, beta_X, and covariate effect, beta_C
beta_X <- -log(1.75)
beta_C <- log(1.5)
lambda <- 0.4
u <- runif(n)
tEVNT <- -log(u) / (lambda * exp(X * beta_X) *exp(C*beta_C))

  # administrative censoring is imposed at 2.5 years
adm_cens <- 2
dEVNT <- as.numeric(tEVNT < adm_cens)
tEVNT <- pmin(tEVNT, adm_cens)

  # data
simdat <- data.frame(ID=ID, X=X, tEVNT=tEVNT, dEVNT=dEVNT, C=C,
                  stringsAsFactors = FALSE)

return(simdat)}

# simulate wide format data
simdat <- fsim_data(seed=123, n=500)
head(simdat)
table(simdat$X, simdat$dEVNT)
```

The simulated dataset 'simdat' consists of the following: a subject identifier (ID), treatment indicator (X), event time (tEVNT), event indicator (dEVNT), covariate (C). Below, the values for the first 6 subjects (out of 500) are shown:

**Sample data:** entries for first 6 subjects in wide format

| ID | X | tEVNT | dEVNT | C |
|---|---|---|---|---|
| 1 | 0 | 0.15207101 | 1 | 0.12439713 |
| 2 | 1 | 0.05414442 | 1 | -0.0618764 |
| 3 | 0 | 1.8411214 | 1 | 0.15608277 |
| 4 | 1 | 2 | 0 | 0.59049665 |
| 5 | 1 | 0.67845763 | 1 | 2.09850877 |
| 6 | 0 | 1.2395858 | 1 | 0.41143489 |

In 'simdat', 167 out of 265 placebo arm patients experienced an event, compared to 106 out of 235 intervention arm patients:

| | **Censored (dEVNT=0)** | **Event (dEVNT=1)** | **Total** |
|---|---|---|---|
| **Control (X=0)** | 98 | 167 | 265 |
| **Intervention (X=1)** | 129 | 106 | 235 |
| **Total** | 217 | 173 | 500 |

The *unadjusted win ratio* can be estimated directly when the data is formatted as above, by applying the winratio function from the WinRatio package.

```
library(WinRatio)

winobj <- winratio(id='ID', trt='X', active='1', data=simdat,
              fu='tEVNT',
              outcomes=list(outc1=c('dEVNT', 's', 'tEVNT')),
              keep.matrix = TRUE)

# saving win/loss/tie matrix for later use
wrmat <- winobj$wr.matrix

# unadjusted win ratio estimate and Z-score
winobj$wr #1.56
winobj$z  #3.46
```

The unadjusted win ratio returns an estimate of 1.56 and Z-value of 3.46.

Prior to estimating the adjusted ordinal win ratio, the data in ‘simdat’ need to be transformed to a long format. For this, we use both ‘simdat’ and the win/loss/tie matrix ‘wrmat’ generated by the winratio command, as shown above. ‘wrmat’ is a matrix of size 235 x 265, where 235 and 265 are the numbers of intervention and control arm patients, respectively. For each pair of patients, a win for the intervention arm patients is denoted 1, a loss -1, and a tie 0.

```
  # Outcome vector Y is obtained using the win/loss tie matrix obtained
  # from the 'winratio' function. We denote losses 0, ties 1, and wins 2.
winmat <- wrmat
winmat[which(wrmat==0)] <- NA; winmat[which(wrmat<0)] <- 0
winmat[which(wrmat>0)] <- 2; winmat[which(is.na(winmat))] <- 1
Y <- as.vector(winmat)
Y <- factor(Y, levels=c(0, 1, 2))

  # Creating the vector of paired differences in covariate C, C_diff
Nc <- table(simdat$X)[1]
Nt <- table(simdat$X)[2]
Cmat_c <- t(matrix(1, nrow=Nc, ncol=Nt)*simdat$C[which(simdat$X==0)])
Cmat_t <- matrix(1, nrow=Nt, ncol=Nc)*simdat$C[which(simdat$X==1)]
C_diff <- as.vector(Cmat_t-Cmat_c)

  # Creating the ID vector for the control group
ID_c <- simdat$ID[which(simdat$X==0)]
matID_c <- matrix(nrow=Nt, ncol=Nc)
for(i in 1:Nc){
  matID_c[1:Nt,i] <- rep(ID_c[i],Nt)}
ID_c <- as.vector(matID_c)

  # Creating the ID vector for the intervention group
ID_t <- simdat$ID[which(simdat$X==1)]
ID_t <- rep(ID_t, Nc)

# long format paired data
simdat_pair <- data.frame(ID_t=ID_t,ID_c=ID_c, Y=Y, C_diff=C_diff)
```

The resulting transformed dataset ‘simdat_pair’ contains 235x265=62275 entries, and consists of two ID variables, one for the intervention group (ID_t) and one for the control group (ID_c), an outcome variable (Y) denoting the win (2), loss (0) and tie (1) status for each pair of patients, when comparing the intervention group patient to the control group patient, and a variable for the difference in covariate C across pairs (C_diff).

**Transformed paired sample data:** entries for first 6 pairs across trial arms

| ID_t | ID_c | Y | C_diff |
|---|---|---|---|
| **2** | 1 | 0 | -0.1862735 |
| **4** | 1 | 2 | 0.46609952 |
| **5** | 1 | 2 | 1.97411164 |
| **7** | 1 | 2 | 1.45640237 |
| **8** | 1 | 2 | 1.00635699 |
| **9** | 1 | 2 | 0.26196298 |

Take, for example, the first row of the transformed paired data above, where intervention group patient with ID_t = 2 is compared to control group patient with ID_c = 1. Patient 2 experiences a loss with respect to patient 1. The table below shows the corresponding, untransformed, wide format data entries for the same patients:

**Unpaired sample data:** wide format entries corresponding to paired entries above

| ID | X | tEVNT | dEVNT | C |
|---|---|---|---|---|
| **1** | 0 | 0.15207101 | 1 | 0.12439713 |
| **2** | 1 | 0.05414442 | 1 | -0.0618764 |
| **4** | 1 | 2 | 0 | 0.59049665 |
| **5** | 1 | 0.67845763 | 1 | 2.09850877 |
| **7** | 1 | 1.23678844 | 1 | 1.5807995 |
| **8** | 1 | 2 | 0 | 1.13075412 |
| **9** | 1 | 2 | 0 | 0.3863601 |

Intervention group patient 2 experiences an event at $t$=0.054, while control group patient 1 experiences an event at $t$=0.152. This is counted as a loss in the paired data, as the intervention group patient has an event before the control group patient. Covariate values are -0.062 and 0.124 for patients 2 and 1, respectively, giving a difference across the pair of C_diff = -0.186. Intervention group patient 4 experiences no event (dEVNT=0) and is censored at $t$=2. When compared to control group patient 1, this is counted as a win in the paired data. And so forth.

The *ordinal adjusted win ratio* is estimated using the transformed paired data. From the model object, we extract the estimated intercepts and transform them to obtain the adjusted loss, win and tie probabilities. From these, we calculate the win ratio and win odds.

```
# performing an ordinal logistic regression on the paired data
ord_adj <- polr(Y ~  C_diff , data = simdat_pair, method='logistic',
            Hess = TRUE)
ord_adj_coefs <- summary(ord_adj)$coefficients

# extracting and transforming the intercept estimates
p_loss_ord_adj <- 1/(1+exp(-ord_adj_coefs[2,1]))
p_win_ord_adj <- 1-1/(1+exp(-ord_adj_coefs[3,1]))
p_tie_ord_adj <- 1-(p_loss_ord_adj+p_win_ord_adj)

# win ratio and win odds
winrat_ord_adj <- p_win_ord_adj/p_loss_ord_adj
winodds_ord_adj <- (p_win_ord_adj+0.5*p_tie_ord_adj)/
             (p_loss_ord_adj+0.5*p_tie_ord_adj)
```

To obtain the approximate standard error for the log win ratio, we extract the model-estimated covariance matrix, using a sandwich estimator to account for both sources of clustering, select the terms corresponding to the intercepts, and apply the delta method. Note that the function *g* is the transformation that is applied to the intercepts to obtain the log win ratio. The means, *x1* and *x2*, are the untransformed intercepts, and *sigma_ord* is the model-estimated covariance matrix for the intercept terms only.

```
# x1 is the untransformed intercept a1 (log odds of Y<=1)
x1 <- summary(ord_adj)$coefficients[3,1]
# x2 is the untransformed intercept a0 (log odds of Y<=0)
x2 <- summary(ord_adj)$coefficients[2,1]

# extracting covariance matrix accounting for the clustering amongst
# the intervention group patients and the control group patients
sigma_ord <- vcovCL(ord_adj, cluster = list(ID_t, ID_c), type = NULL,
              sandwich = TRUE)
```

```
# selecting terms relating to intercepts and reordering rows so that
# row 1 corresponds to x1 and row 2 to x2
sigma_ord <- sigma_ord[2:3,2:3]
sigma_ord <- sigma_ord[c(2,1),c(2,1)]

# applying the deltamethod function from the msm package
library(msm)
logord_winrat_SE <-
  deltamethod(g= ~ log((1-(1/(1+exp(-x1))))/(1/(1+exp(-x2)))),
            mean = c(x1,x2),
            cov = sigma_ord,
            ses = TRUE)
```

From the log win ratio SE we obtain the Z-score and the 95% CI:

```
# log ordinal WR SE =  0.134
logord_winrat_SE
# ordinal WR = 1.733
winrat_ord_adj
# Z-score = 4.099
Z_ord_winrat <- log(winrat_ord_adj)/logord_winrat_SE
# 95% CI = 1.332 to 2.254
CI_ord_winrat <- exp(c(log(winrat_ord_adj)-qnorm(0.975)*logord_winrat_SE,
                  log(winrat_ord_adj)+qnorm(0.975)*logord_winrat_SE))
```

Recall that the unadjusted win ratio returned an estimate of 1.56 and Z-value of 3.46. The ordinal adjusted win ratio returns an estimate of 1.733 and Z-value of 4.10.

Note that using the deltamethod function from the msm package is equivalent to performing the manual calculation described in Section A.1:

```
# taking the partial derivative of log((1-(1/(1+exp(-x1))))/
                              (1/(1+exp(-x2)))):
# with respect to x1 and with respect to x2:
deriv_wrt_x1 <- -1/(exp(-x1)+1)
deriv_wrt_x2 <- -1/(exp(x2)*(1+exp(-x2)))
deriv_vec <- c(deriv_wrt_x1, deriv_wrt_x2)

# obtaining log win ratio SE
sqrt(deriv_vec%*%sigma_ord%*%deriv_vec) # 0.134
```

#### Appendix B: Adaptation of the randomisation-based method to calculate the adjusted win ratio and win difference

*In this appendix we describe our adaptation of the randomisation-based method for covariate adjustment, which estimates the adjusted Mann-Whitney probability and the win odds, to the win ratio. In Section B.1, we derive an approximate standard error, in Section B.2 we illustrate the properties of the adjusted win ratio estimator and its SE in simulation, and in Section B.3 we describe how multiple covariates can be adjusted for. Section B.4 gives a worked example for estimating the adjusted win ratio with approximate SE.*

Since the win ratio cannot be calculated directly using the randomisation-based method, we use an equivalent methodology to estimate an adjusted win ratio and win difference as follows. The methodology is analogous to the method described by Gasparyan, except that ties are not directly used in the various calculations. We first obtain the unadjusted win probability, $\hat{\vartheta}$, which is equal to P(win)=$\frac{\#wins}{\#pairwise\ comparisons}$, and the unadjusted loss probability, $\hat{\lambda}$, which is equal to P(loss)= $\frac{\#losses}{\#pairwise\ comparisons}$. The adjusted win probability, $\widehat{\vartheta_a}$, is obtained as in (1) but now with *p* and *q* replaced by *u* and *v*, where *u* represents the win probabilities for individuals in the intervention arm when compared to the control arm, and *v* the win probabilities for individuals in the control arm when compared to the intervention arm. For a patient *i* in the intervention arm, the individual win probability, $u_i$, is given by

$$u_i = \frac{1}{N_C}\sum_{j=1}^{N_C} K_{ij},$$

where $K_{ij} = 1$ if the $i$th subject from the intervention arm wins against the $j$th subject from the control arm, and 0 otherwise.

For a patient *j* in the control arm, the individual win probability, $v_j$, is given by

$$v_j = \frac{1}{N_I}\sum_{i=1}^{N_I} L_{ij},$$

where $L_{ij} = 1$ if $i$th subject from the intervention arm loses against the $j$th subject from the control arm (or, equivalently, if the *j*th subject from the control arm wins against the *i*th subject from the intervention arm) and 0 otherwise.

The adjusted win probability, $\widehat{\vartheta_a}$, is then obtained by

$$\widehat{\vartheta_a} = \hat{\vartheta} - \frac{\overline{C_I} - \overline{C_C}}{\frac{\text{var}(C_I)}{N_I} + \frac{\text{var}(C_C)}{N_C}} \left[ \frac{\text{Cov}(C_I, u)}{N_I} + \frac{\text{Cov}(C_C, v)}{N_C} \right], \quad (3)$$

with all remaining terms defined as in equation (2) in the main text. That is, $C_I$ and $C_C$ *r*epresent the covariate values for individuals in the intervention and control arm respectively, and $N_I$ and $N_C$ are the number of patients in the two groups.

The adjusted loss probability, $\widehat{\lambda_a}$, is obtained analogously, but now using $u^-$ and $v^-$, which represent the loss probabilities for individuals in the intervention arm when compared to the control arm, and individuals in the control arm, when compared to the intervention arm, respectively. For example, for a patient *i* in the intervention arm, the individual loss probability, $u_i^-$, is given by

$$u_i^-, = \frac{1}{N_C} \sum_{j=1}^{N_C} L_{ij}.$$

The ratio and the difference of the adjusted win and loss probability will give us the adjusted win ratio and adjusted win difference, respectively. Note that, unlike for the adjusted win odds, we must separately estimate both the adjusted win probability and loss probability. The adjusted loss probability cannot be inferred from the adjusted win probability, as this requires knowing the tie probability, which may also be affected by the adjustment.

The adjusted tie probability, $\widehat{\tau_a}$, is obtained analogously to the adjusted win and loss probabilities. For a patient *i* in the intervention arm, the individual tie probability, $s$, is given by

$$s_i = \frac{1}{N_C} \sum_{j=1}^{N_C} T_{ij},$$

where $T_{ij} = 1$ if $i$th subject from the intervention and the $j$th subject from the control arm are tied. For a patient *j* in the control arm, the individual tie probability, $t_i$, is given by

$$t_i = \frac{1}{N_I}\sum_{j=1}^{N_I} T_{ij},$$

The adjusted tie probability, $\widehat{\tau_a}$, is then obtained as in (2), replacing *u* with *s* and *v* with *t.* Note that the adjusted win, loss and tie probabilities, $\widehat{\vartheta_a}$, $\widehat{\lambda_a}$, and $\widehat{\tau_a}$, add up to 1, just as the unadjusted probabilities add up to 1.

#### B.1 Deriving an SE for the adjusted win ratio

To obtain a Z-value for the covariate-adjusted win ratio, $Z_{\log(\mathrm{WR}_\alpha)} = \frac{\log(\mathrm{WR}_\alpha)}{\mathrm{SE}_{\log(\mathrm{WR}_\alpha)}}$, we require the standard error of the log adjusted win ratio estimate. Gasparyan *et al.* (…) derived the applicable squared standard error for the Mann-Whitney (win+0.5 x tie) probability. We derive analogous squared standard errors for the adjusted win probability, $\sigma^2_{\vartheta_\alpha}$, adjusted loss probability, $\sigma^2_{\lambda_\alpha}$, adjusted tie probability, $\sigma^2_{\tau_\alpha}$:

$$\sigma^2_{\vartheta_\alpha} = \frac{\mathrm{var}(u)}{N_I} + \frac{\mathrm{var}(v) + \mathrm{var}(t) + 2\mathrm{cov}(v,t)}{N_C} - \left[\frac{\left(\frac{\mathrm{Cov}(C_I,u)}{N_I} + \frac{\mathrm{Cov}(C_C,v) + \mathrm{Cov}(C_C,t)}{N_C}\right)^2}{\frac{\mathrm{var}(C_I)}{N_I} + \frac{\mathrm{var}(C_C)}{N_C}}\right]$$

$$\sigma^2_{\lambda_\alpha} = \frac{\mathrm{var}(u^-)}{N_I} + \frac{\mathrm{var}(v^-) + \mathrm{var}(t) + 2\mathrm{cov}(v^-,t)}{N_C} - \left[\frac{\left(\frac{\mathrm{Cov}(C_I,u^-)}{N_I} + \frac{\mathrm{Cov}(C_C,v^-) + \mathrm{Cov}(C_C,t)}{N_C}\right)^2}{\frac{\mathrm{var}(C_I)}{N_I} + \frac{\mathrm{var}(C_C)}{N_C}}\right]$$

$$\sigma^2_{\tau_\alpha} = \frac{\text{var}(s)}{N_I} + \frac{\text{var}(t))}{N_C} - \left[\frac{\left(\frac{\text{Cov}(C_I, s)}{N_I} + \frac{\text{Cov}(C_C, t)}{N_C}\right)^2}{\frac{\text{var}(C_I)}{N_I} + \frac{\text{var}(C_C)}{N_C}}\right]$$

From these, using standard variance properties, we can calculate the covariance between the adjusted win and loss probabilities, $\sigma_{\vartheta_\alpha\lambda_\alpha}$:

$$\text{var}(\tau_\alpha) = \text{var}(1 - \tau_\alpha) = \text{var}(\vartheta_\alpha + \lambda_\alpha) = \text{var}(\vartheta_\alpha) + \text{var}(\lambda_\alpha) + 2\,\text{cov}(\vartheta_\alpha, \lambda_\alpha).$$

Then

$$\sigma_{\vartheta_\alpha\lambda_\alpha} = \text{cov}(\vartheta_\alpha, \lambda_\alpha) = \frac{\text{var}(\tau_\alpha) - \text{var}(\vartheta_\alpha) - \text{var}(\lambda_\alpha)}{2}.$$

Let $\Sigma$ denote the covariance matrix for the adjusted win and loss probabilities:

$$\Sigma = \begin{bmatrix} \sigma^2_{\vartheta_\alpha} & \sigma_{\vartheta_\alpha\lambda_\alpha} \\ \sigma_{\vartheta_\alpha\lambda_\alpha} & \sigma^2_{\lambda_\alpha} \end{bmatrix}.$$

With $\Sigma$, we obtain $\text{SE}_{\log(\text{WR}_\alpha)}$ using the delta method. Let $f(\vartheta_\alpha, \lambda_\alpha) = \log\left(\frac{\vartheta_\alpha}{\lambda_\alpha}\right)$, which estimates the adjusted log win ratio. The corresponding variance, $\text{var}(f(\vartheta_\alpha, \lambda_\alpha))$ = $\text{var}[\log(\text{WR}_\alpha)]$, is then calculated using the first order derivatives of $f(\vartheta_\alpha, \lambda_\alpha)$ with respect to $\vartheta_\alpha$ and $\lambda_\alpha$, and the covariance matrix, $\Sigma$:

$$\text{var}[\log(\text{WR}_\alpha)] \approx \begin{bmatrix} \frac{\partial f}{\partial \vartheta_\alpha} & \frac{\partial f}{\partial \lambda_\alpha} \end{bmatrix} \times \Sigma \times \begin{bmatrix} \frac{\partial f}{\partial \vartheta_\alpha} & \frac{\partial f}{\partial \lambda_\alpha} \end{bmatrix} = \begin{bmatrix} \frac{1}{\vartheta_\alpha} & \frac{-1}{\lambda_\alpha} \end{bmatrix} \times \Sigma \times \begin{bmatrix} \frac{1}{\vartheta_\alpha} & \frac{-1}{\lambda_\alpha} \end{bmatrix}.$$

In the section below, we show in simulation that this approximate SE for the log adjusted win ratio gives near identical results as the Monte Carlo-derived standard error.

#### B.2 Estimator properties in simulation

We investigated the properties of the RB-adapted win ratio estimator in simulation, for a composite time-to-event outcome of death and hospitalisation, and a single prognostic baseline covariate, log NT-proBNP, employing the same data-generating mechanism as described in Section 4.1 of the main text. Estimates were obtained when an effect of treatment was present, when adjusting for a prognostic or non-prognostic covariate, and under the null. SEs were estimated by taking the Monte Carlo standard deviation across the log win ratio estimates and by calculating approximate analytical SEs as per Section B.1. Results are shown in the table below.

**Appendix B Table:** Randomisation-based win ratio estimator properties in simulation, for a hierarchical composite outcome of time to death or hospitalization, with treatment effect and under the null, when adjusting for a prognostic or non-prognostic covariate.

| | | | **Using Monte Carlo SD** | | | **Using analytical SE** | | |
|---|---|---|---|---|---|---|---|---|
| | Mean win ratio | Mean log win ratio | Log win ratio SE (MCSD) | Mean Z-statistic | Power | Mean log win ratio SE (analytical) | Mean Z-statistic | Power/ type-1 error |
| *With treatment effect and prognostic covariate (HR=1.75 per 1 SD increase)* | | | | | | | | |
| **Unadjusted** | 1.471 | 0.378 | 0.125 | 3.033 | 0.861 | 0.126 | 3 | 0.859 |
| **Randomisation-based** | 1.469 | 0.378 | 0.117 | 3.239 | 0.897 | 0.118 | 3.184 | 0.896 |
| *With treatment effect and non-prognostic covariate* | | | | | | | | |
| **Unadjusted** | 1.526 | 0.415 | 0.127 | 3.270 | 0.905 | 0.127 | 3.262 | 0.908 |
| **Randomisation-based** | 1.526 | 0.415 | 0.127 | 3.268 | 0.904 | 0.127 | 3.265 | 0.909 |
| *With null treatment effect and prognostic covariate (HR=1.75 per 1 SD increase)* | | | | | | | | |
| **Unadjusted** | 1.008 | 0.001 | 0.117 | 0.012 | 0.025 | 0.118 | 0.012 | 0.023 |
| **Randomisation-based** | 1.007 | 0.001 | 0.109 | 0.010 | 0.025 | 0.11 | 0.01 | 0.024 |

Adjusting for a prognostic covariate resulted in a decreased SE (0.117 compared to 0.125) and a corresponding increase in power, while the point estimate changed very little (1.469 compared to 1.471), which is expected when estimating a marginal treatment effect. Adjusting for a non-prognostic covariate gave very similar results to the unadjusted estimator, with point estimates and SEs equal when rounded to three decimal places, and minimal differences in Z-statistics and power. Under the null, the two-sided type-1 error rate was 0.025. Results were similar when using the Monte-Carlo derived SE and when using the analytical SE.

#### B.3 Adjusting for multiple covariates

Both the randomisation-based method, as described by Gasparyan *et al.* for the adjusted Mann-Whitney probability and our extended approach for the adjusted win ratio generalise to multiple covariates. Each additional covariate is accounted for by a separate adjustment, in which any collinearity between predictors is taken into account. A parallel can be drawn with performing multiple simple linear regression versus a single multiple linear regression. First, consider once more the case where we adjust for a single covariate only. The covariate-adjusted win probability is given in (2). Here, the mean difference of the covariate across trial arms is multiplied by the covariance of the covariate with the individual win probabilities in the intervention group and control group, and divided by the covariate variances, with all terms weighted by group size. An equivalent result can be achieved by performing a weighted linear regression of the combined vector of win probabilities, $[u, v]$, on a treatment group indicator, X (where X=1 if the individual is in the intervention arm), and covariate, C:

$$[u, v] \sim \alpha + \beta X + \gamma_1 C + \varepsilon \,, \tag{4}$$

with weights *W,* where $W = \frac{N_I}{N_C}$ if *X*=0, and $W = \frac{N_C}{N_I}$ if *X*=1. The adjusted win probability, is then given by

$$\widehat{\vartheta_a} = \hat{\vartheta} - (\bar{C_I} - \bar{C}_C) \times \gamma_1 \tag{5}$$

with (5) giving an estimate identical to (2). When adjusting also for a second covariate, Z, the above process is repeated, but now, in addition to *C* we also adjust for Z*,* and equation (4) changes to:

$$[u, v] \sim \alpha + \beta X + \delta_1 C + \ \delta_2 Z + \varepsilon \,. \tag{6}$$

The win probability adjusted for both covariates, *C* and *Z*, is then given by

$$\widehat{\vartheta_{am2}} = -(\bar{C_I} - \bar{C}_C) \times \delta_1 - \ (\bar{Z_I} - \bar{Z}_C) \times \ \delta_2 \,. \tag{7}$$

The corresponding algebraic expression for the win probability when adjusting for covariates *C* and *Z* simultaneously is given below and becomes increasingly cumbersome the more covariates are included. Therefore we recommend, when adjusting for multiple covariates, employing the linear regression approach using equations (6) and (7), over applying the algebraic correction.

$$\widehat{\vartheta_{am2}} = \hat{\vartheta} - \left\{ \begin{array}{c} (\overline{C_I} - \overline{C_C}) \times \left[ \begin{array}{c} \left(\frac{\text{Cov}(C_I,u)}{N_I} + \frac{\text{Cov}(C_C,v)}{N_C}\right) \times \left(\frac{\text{var}(Z_I) +}{N_I} + \frac{\text{var}(Z_C)}{N_C}\right) - \\ \left(\frac{\text{Cov}(Z_I,u)}{N_I} + \frac{\text{Cov}(Z_C,v)}{N_C}\right) \times \left(\frac{\text{cov}(C_I,Z_I\ ) +}{N_I} + \frac{\text{cov}(C_C,Z_C)}{N_C}\right) \end{array} \right] + \\ (\overline{Z_I} - \overline{Z_C}) \times \left[ \begin{array}{c} \left(\frac{\text{Cov}(Z_I,u)}{N_I} + \frac{\text{Cov}(Z_C,v)}{N_C}\right) \times \left(\frac{\text{var}(C_I) +}{N_I} + \frac{\text{var}(C_C)}{N_C}\right) - \\ \left(\frac{\text{Cov}(C_I,u)}{N_I} + \frac{\text{Cov}(C_C,v)}{N_C}\right) \times \left(\frac{\text{cov}(C_I,Z_I\ ) +}{N_I} + \frac{\text{cov}(C_C,Z_C)}{N_C}\right) \end{array} \right] \end{array} \right\} \times$$

$$\frac{1}{\left(\frac{\text{cov}(C_I,Z_I\ ) +}{N_I} + \frac{\text{cov}(C_C,Z_C)}{N_C}\right)^2 + \left(\frac{\text{var}(C_I) +}{N_I} + \frac{\text{var}(C_C)}{N_C}\right) \times \left(\frac{\text{var}(Z_I) +}{N_I} + \frac{\text{var}(Z_C)}{N_C}\right)}$$

#### B.4. Worked example

First, we simulate a sample dataset using the function fsim_data, as described in Appendix A.3 (Worked example for ordinal win ratio estimator). This will generate a wide format dataframe *simdat* with a single time-to-event outcome (tEVNT), an event indicator (dEVNT), treatment group indicator (X=[0,1] for control and intervention group, respectively, and a covariate vector (C). To apply the randomisation-based estimator, we first obtain the relevant unadjusted quantities using the conventional win ratio estimator from the WinRatio package, and extract the win/loss/tie matrix from the resulting object.

```
library(WinRatio)
  # generating data
simdat <- fsim_data(seed=123, n=500)

  # obtaining unadjusted win ratio object
winobj <- winratio(id='ID', trt='X', active='1', data=simdat, fu='tEVNT',
              outcomes=list(outc1=c('dEVNT', 's', 'tEVNT')),
              keep.matrix = TRUE)

  # extracting win/loss/tie matrix
wrmat <- winobj$wr.matrix

  # unadjusted win, loss, tie win+0.5xtie probabilities
package_winprop <- winobj$total.wins/(winobj$n1*winobj$n0) #0.485
package_lossprop <- winobj$total.loss/(winobj$n1*winobj$n0) #0.312
package_tieprop <-  winobj$total.ties/(winobj$n1*winobj$n0) #0.203
```

```
package_wintieprob <- (winobj$total.wins + 0.5*winobj$total.ties)/
                    (winobj$n1*winobj$n0) #0.587

  # unadjusted win ratio, win odds, and Z-value
package_winrat <- package_winprop/package_lossprop #1.556
package_winodds <- package_wintieprob/(1-package_wintieprob) #1.419
winobj$wr #1.556
winobj$z  #3.461
```

This yields an unadjusted win ratio and win odds estimates of 1.56 and 1.42, respectively, with a Z-value of 3.46.

We use the win/tie/loss matrix *wrmat* to obtain the vectors of the individual win+0.5xtie, win, loss, and tie probabilities for each intervention group. First, we obtain the vectors of the individual win+0.5xtie probabilities for each intervention group, and use these to calculate the adjusted win+0.5xtie probability and resulting odds ratio, using the original randomisation-based estimator. Note we define weights *wts* for the linear regression (see also Appendix B.3, eq. 3).

```
# ps: individual win + tie probabilities for the intervention group wrt
# the control group
ps <- apply(wrmat, 1, function(x){
  return((length(which(x>0))+0.5*length(which(x==0)))/length(x))})
# qs: individual win + tie probabilities for the control group wrt the
# intervention group
qs <- apply(wrmat, 2, function(x){
  return((length(which(x<0)) + 0.5*length(which(x==0))) /length(x))})
pqs <- c(qs, ps)

simdatI <- simdat[which(simdat$x==1),]
simdatC<- simdat[which(simdat$x==0),]

# vector of covariate values
cs <- c(simdatC$C, simdatI$C)
# treatment indicator
trs <- c(simdatC$x, simdatI$x)

# group sizes (nI and nC for intervention and control group, resp.)
nI <- length(which(simdat$x==1)); nC <- length(which(simdat$x==0))
```

```
# weights according to intervention group size
wts <- c(rep(nI/nC, nC), rep(nC/nI, nI))

# obtaining adjusted win+0.5*tie probability and win odds
winpropties_lin_adj <- package_wintieprob -
  summary(lm(cs~trs))$coefficients[2,1]*
  summary(lm(pqs~trs+cs, weights=wts))$coefficients[3,1] #0.596
winodds_lin_adj <- winpropties_lin_adj/(1-winpropties_lin_adj) #1.475121
```

This gives an adjusted win odds estimate of 1.48.

Now we obtain the adjusted win, loss, and tie probabilities per our adapted approach. First we obtain the individual win, loss and tie probabilities for each group:

```
# us: individual win probabilities for the intervention group
# wrt the control group
us <- apply(wrmat, 1, function(x){
  return(length(which(x>0))/length(x))})
# us: individual loss probabilities for the intervention group
# wrt the control group
usi <- apply(wrmat, 1, function(x){
  return(length(which(x<0))/length(x))})
# vs: individual win probabilities for the control group
# wrt the intervention group .. how many losses did c pat have
vs <- apply(wrmat, 2, function(x){
  return(length(which(x<0))/length(x))})
# vs: individual loss probabilities for the control group
# wrt the intervention group
vsi <- apply(wrmat, 2, function(x){
  return(length(which(x>0))/length(x))})
# s: individual tie probabilities for the intervention group
# wrt the control group
s <- apply(wrmat, 1, function(x){
  return(length(which(x==0))/length(x))})
# t: individual tie probabilities for the control group
# wrt the intervention group
t <- apply(wrmat, 2, function(x){
  return(length(which(x==0))/length(x))})

# vectors of win probabilities (vus), loss probabilities (vusi),
```

```
# and tie probabilites (ts)
vus <- c(vs, us)
vusi <- c(vsi, usi)
ts <- c(t, s)
```

Using *vus* (vector of individual win probabilities), *vusi* (vector of individual loss probabilities), and *ts* (vector of individual tie probabilities), we obtain the adjusted win, loss, and tie probabilities. We calculate the adjusted win ratio from the adjusted win and loss probabilities, and the adjusted win odds from the adjusted win, loss and tie probabilities.

```
# obtaining adjusted win probability, loss probability, and tie probability
winpr_lin_adj <- package_winprop - summary(lm(cs~trs))$coefficients[2,1]*
              summary(lm(vus~trs+cs, weights=wts))$coefficients[3,1]
losspr_lin_adj<- package_lossprop - summary(lm(cs~trs))$coefficients[2,1]*
              summary(lm(vusi~trs+cs, weights=wts))$coefficients[3,1]
tiepr_lin_adj <- package_tieprop - summary(lm(cs~trs))$coefficients[2,1]*
              summary(lm(ts~trs+cs, weights=wts))$coefficients[3,1]

# adjusted win (0.4906), tie (0.2108), and loss (0.2986) probabilities all
# add up to 1:
winpr_lin_adj+losspr_lin_adj+tiepr_lin_adj #1

# adjusted win ratio
winrat_lin_adj <- winpr_lin_adj/losspr_lin_adj #1.642

# calculating adjusted win odds from win, loss and tie probabilities instead
# of the win+0.5*tie probability per the original randomisation-based
# method: 1.475121
(winpr_lin_adj+0.5*tiepr_lin_adj)/(losspr_lin_adj+0.5*tiepr_lin_adj)
```

The individual adjusted probabilities are 0.491, 0.299 and 0.211 for the win, loss and tie probability, respectively, and add up to 1.  The adjusted win ratio and win odds are 1.64 and 1.48, respectively. Note that the win odds, here calculated from the adjusted win, loss and tie probabilities, is identical (within 6 decimals) to the estimate obtained when using the win+0.5xtie probability, when employing the original method.

To calculate an approximate SE for the log adjusted win ratio, we need the squared SEs for the adjusted win probability, adjusted loss probability and their covariance. To obtain the latter, we additionally need the squared SE for the adjusted tie probability. (see also algebraic notation in Appendix B1)

```
# covariate vectors per group (split out for ease of notation)
CC <- simdatC$C
CI <- simdatI$C

# win probability SE
winprSE <- sqrt(var(us)/nI+var(vs)/nC+var(t)/nC+2*cov(vs,t)/nC-
                  ((cov(vs, CC)/nC+cov(us, CI)/nI+cov(t, CC)/nC)^2/
                    (var(CC)/nC+var(CI)/nI)))

# loss probability SE
lossprSE <- sqrt(var(usi)/nI+var(vsi)/nC+var(t)/nC+2*cov(vsi,t)/nC-
                  ((cov(vsi, CC)/nC+cov(usi, CI)/nI+cov(t, CC)/nC)^2/
                    (var(CC)/nC+var(CI)/nI)))

# tie probability SE
tieprSE <- sqrt(var(s)/nI+var(t)/nC-
                 ((cov(t, CC)/nC+cov(s, CI)/nI)^2/
                   (var(CC)/nC+var(CI)/nI)))

# win/loss probability covariance
cov_winloss <- (tieprSE^2 - winprSE^2 - lossprSE^2)/2
```

We apply the delta method using the *deltamethod* function form the msm package to obtain the SE for the log adjusted win ratio:

```
x1 <- winprop_lin_adj
x2 <- lossrop_lin_adj
covmat <- matrix(c(winprSE ^2, cov_winloss, cov_winloss, lossprSE ^2),
                 nrow=2,ncol=2)
library(msm)
log_lin_adjwinrat_SE <- deltamethod(g= ~ log(x1/x2),
                                    mean = c(x1,x2),
                                    cov = covmat,
                                    ses = TRUE)
```

From the log win ratio SE we obtain the Z-score and the 95% CI:

```
# log WR SE =  0.1268
logadjwinrat_SE
# adjusted WR = 1.733
winrat_lin_adj
# Z-score = 3.914
Z_lin_winrat <- log(winrat_lin_adj)/log_lin_adjwinrat_SE
# 95% CI = 1.281 to 2.107
CI_lin_winrat <- exp(c(
        log(winrat_lin_adj)-qnorm(0.975)*log_lin_adjwinrat_SE,
        log(winrat_lin_adj)+qnorm(0.975)*log_lin_adjwinrat_SE))

# unadjusted WR Z-score:
winobj$z #3.461
```

The Z-score for the adjusted win ratio is 3.91 compared to 3.46 for the unadjusted WR.

*A note on adjusting for multiple covariates:*

When adjusting for multiple covariates, an adjustment term needs to be specified for each covariate, while taking into account the correlations between the covariates. See also equations (5) and (6) in appendix B3; this is done by including both covariates in the second regression. E.g., the adjusted win probability would be estimated as follows:

```
# adjusting for a single covariate (vector ‘cs’)
winpr_lin_adj <- package_winprop - summary(lm(cs~trs))$coefficients[2,1]*
        summary(lm(vus~trs+cs, weights=wts))$coefficients[3,1]
# adjusting for two covariates (vectors ‘cs’ and ‘cs2’): requires two
# adjustment terms, with both covariates included in the second regression
winpr_lin_adj_mult <- package_winprop –
        summary(lm(cs~trs))$coefficients[2,1]*
      summary(lm(vus~trs+cs+cs2, weights=wts))$coefficients[3,1] –
        summary(lm(cs2~trs))$coefficients[2,1]*
      summary(lm(vus~trs+cs +cs2, weights=wts))$coefficients[4,1] –
```

### Appendix C: Calculation of effective increase in sample size based on Z-statistic from simulations

Let $Z_u$ denote the median Z-statistic for the unadjusted estimator, which can be defined as the median standardised unadjusted log treatment effect, and $Z_a$ the corresponding quantity for the adjusted estimator (i.e. the median standardised adjusted log treatment effect). The effective increase in sample size is then given by $\left[\left(\frac{Z_a}{Z_u}\right)^2 - 1\right] * 100\%$.
